# Circulating proteins to predict adverse COVID-19 outcomes

**DOI:** 10.1101/2021.10.04.21264015

**Authors:** Chen-Yang Su, Sirui Zhou, Edgar Gonzalez-Kozlova, Guillaume Butler-Laporte, Elsa Brunet-Ratnasingham, Tomoko Nakanishi, Wonseok Jeon, David Morrison, Laetitia Laurent, Jonathan Afilalo, Marc Afilalo, Danielle Henry, Yiheng Chen, Julia Carrasco-Zanini, Yossi Farjoun, Maik Pietzner, Nofar Kimchi, Zaman Afrasiabi, Nardin Rezk, Meriem Bouab, Louis Petitjean, Charlotte Guzman, Xiaoqing Xue, Chris Tselios, Branka Vulesevic, Olumide Adeleye, Tala Abdullah, Noor Almamlouk, Yara Moussa, Chantal DeLuca, Naomi Duggan, Erwin Schurr, Nathalie Brassard, Madeleine Durand, Diane Marie Del Valle, Ryan Thompson, Mario A. Cedillo, Eric Schadt, Kai Nie, Nicole W Simons, Konstantinos Mouskas, Nicolas Zaki, Manishkumar Patel, Hui Xie, Jocelyn Harris, Robert Marvin, Esther Cheng, Kevin Tuballes, Kimberly Argueta, Ieisha Scott, The Mount Sinai COVID-19 Biobank Team, Celia M T Greenwood, Clare Paterson, Michael A. Hinterberg, Claudia Langenberg, Vincenzo Forgetta, Joelle Pineau, Vincent Mooser, Thomas Marron, Noam Beckmann, Ephraim Kenigsberg, Seunghee Kim-schulze, Alexander W. Charney, Sacha Gnjatic, Daniel E. Kaufmann, Miriam Merad, J Brent Richards

## Abstract

Predicting COVID-19 severity is difficult, and the biological pathways involved are not fully understood. To approach this problem, we measured 4,701 circulating human protein abundances in two independent cohorts totaling 986 individuals. We then trained prediction models including protein abundances and clinical risk factors to predict adverse COVID-19 outcomes in 417 subjects and tested these models in a separate cohort of 569 individuals. For severe COVID-19, a baseline model including age and sex provided an area under the receiver operator curve (AUC) of 65% in the test cohort. Selecting 92 proteins from the 4,701 unique protein abundances improved the AUC to 88% in the training cohort, which remained relatively stable in the testing cohort at 86%, suggesting good generalizability. Proteins selected from different adverse COVID-19 outcomes were enriched for cytokine and cytokine receptors, but more than half of the enriched pathways were not immune-related. Taken together, these findings suggest that circulating proteins measured at early stages of disease progression are reasonably accurate predictors of adverse COVID-19 outcomes. Further research is needed to understand how to incorporate protein measurement into clinical care.

## Introduction

A remarkable feature of COVID-19 disease is its highly variable clinical course, where some individuals manifest severe disease or death, and others remain asymptomatic. Several clinical and genetic risk factors explain a proportion of these outcomes^1–5^, yet most of the host biological causes of these adverse COVID-19 outcomes remain unknown.

Recent reports have identified some of the biologic pathways influencing risk of adverse COVID-19, such as immune responses^6–9^, interferon pathways^10–12^, and T-cell dysfunction^13,14^. However, many such studies have focused on narrow sets of pre-selected cytokines. One way to rapidly assess thousands of potential biomarkers associated with the severity of COVID-19 is through the measurement of blood circulating proteins. Such circulating proteins may be useful because they can help to identify pathways influencing severity of disease. They may also identify individuals at high risk of a severe COVID-19 clinical course. Similarly, circulating proteomic biomarkers have recently been shown to serve as predictors of other common diseases^15–21^ including cardiovascular disease. They are also relevant in drug discovery because they are generally more accessible to pharmacological manipulation than intracellular proteins^22–26^. Thus, understanding the circulating proteins associated with adverse COVID-19 outcomes may be helpful to address major challenges raised by the current pandemic^13,27–38^.

We undertook a large-scale study to assess the relationship of thousands of circulating proteins with COVID-19 outcomes. To do so, we used machine learning methods to develop a predictive model of COVID-19 severity using the circulating blood protein abundances as predictors. Proteins were measured using 4,984 nucleic acid aptamers (SOMAmer reagents)^39^ targeting 4,701 unique circulating human proteins in two cohorts collected from two countries, which in total included 986 individuals. The training cohort was comprised of 417 individuals from two sites of the Biobanque Québécoise de la COVID-19 (BQC19 cohort). This cohort was used to train a model to predict adverse COVID-19 outcomes. This model was then tested in a separate test cohort from the Mount Sinai Hospital in New York City, which was similarly characterized for the same protein measurements and COVID-19 outcomes.

This large-scale study across two countries and two geographically separated cohorts identified circulating proteins associated with COVID-19 outcomes measured at a large-scale in well-characterized cohorts. These findings provide insights into the biological pathways influencing these outcomes and the ability of proteomics to predict these outcomes.

## Results

### Cohorts

To establish a proteomic-based prediction model for adverse COVID-19, we used the BQC19 cohort, which consisted of samples from two hospitals in Montreal, with proteomic measurements for training and cross-validation. The final model was tested in an independent cohort from Mount Sinai hospital in New York City. Using the same SomaLogic® assay, 4,984 SOMAmer reagents measured the levels of 4,701 different circulating proteins in both the BQC19 and Mount Sinai cohorts. To train our models, we selected 417 individuals which included 313 nasal swab SARS-CoV-2 PCR positive patients with baseline samples collected within 14 days of symptom onset (mean and median time since symptom onset in COVID-19 patients = 7.0 days (SD = 3.96 days)). The BQC19 cohort also included an additional 104 individuals who presented to the same hospital sites with symptoms consistent with COVID-19 but had a negative SARS-CoV-2 PCR nasal swab. The Mount Sinai cohort consisted of 569 individuals with their earliest samples also collected within 14 days of symptom onset. Among them were 472 SARS-CoV-2 positive patients again confirmed by PCR, one patient confirmed by chest CT, and 96 SARS-CoV-2 negative individuals (89 with PCR confirmation). If multiple blood samples were collected from the same person, we used the samples collected at the time point closest to symptom onset. We chose to use samples close to symptom onset to reflect the proteome of acute COVID-19, rather than its recovery phase.

The demographic and clinical characteristics of the participants in the training and testing datasets are shown in **Table 1**. In the BQC19 cohort, the mean age across all samples was 65.3 years (SD = 18.4 years), and 52% of the cohort were men. In the Mount Sinai cohort, the mean age was 59.6 years (SD: 19.4 years), and 58.2% of the cohort were men.

**Table 1.**
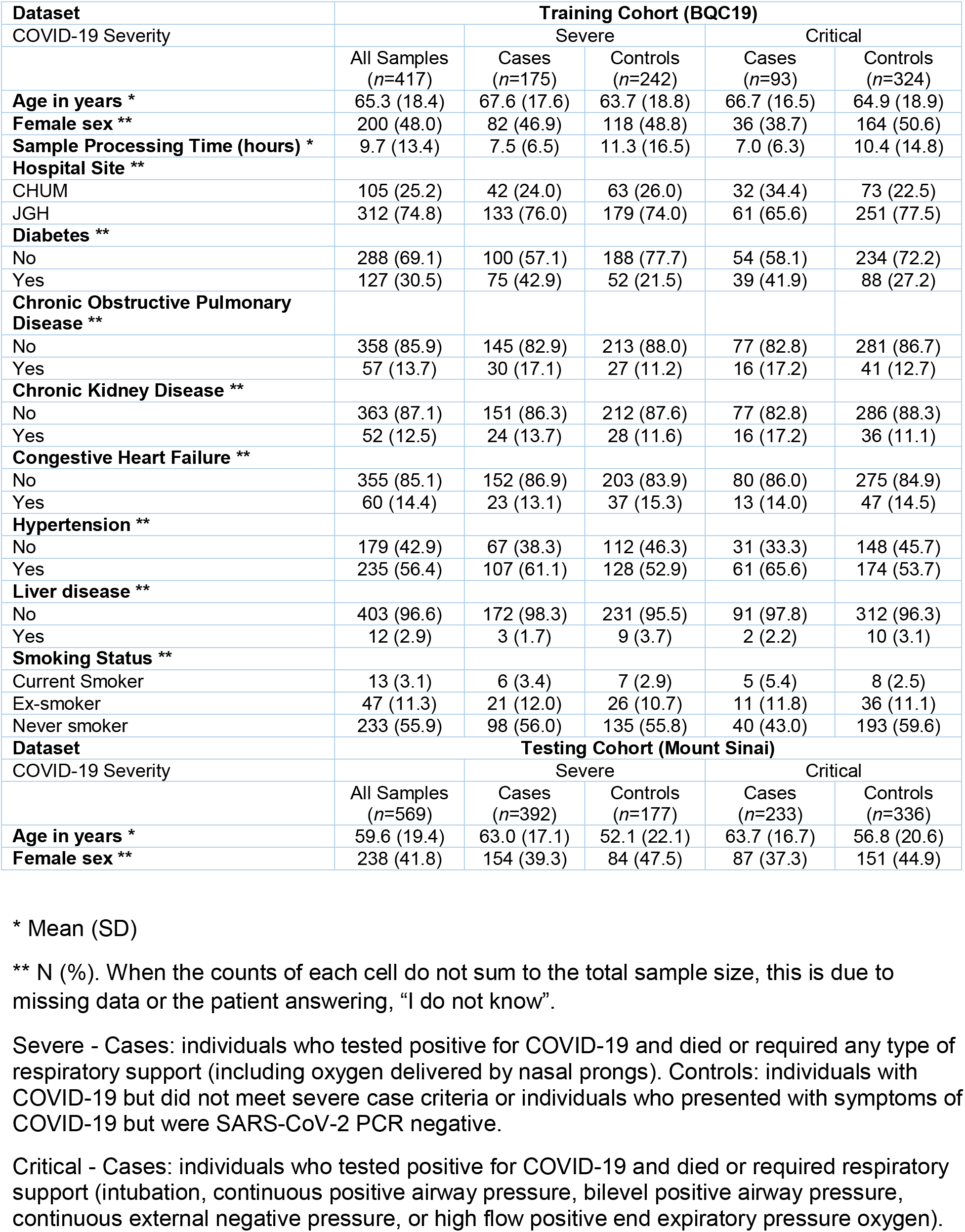
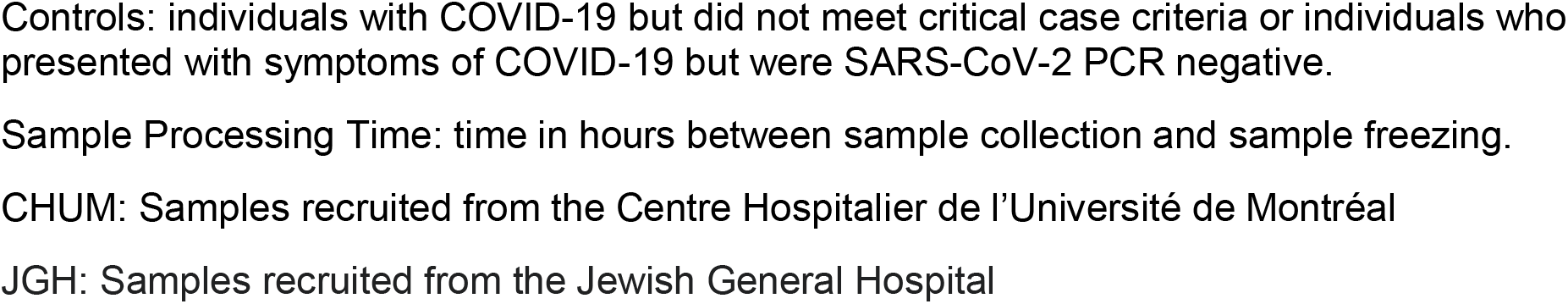
Demographic characteristics of the participating cohorts.

| **Dataset** | **Training Cohort (BQC19)** |  |  |  |  |
| --- | --- | --- | --- | --- | --- |
| COVID-19 Severity |  | Severe |  | Critical |  |
|  | All Samples<br>(n=417) | Cases<br>(n=175) | Controls<br>(n=242) | Cases<br>(n=93) | Controls<br>(n=324) |
| **Age in years \*** | 65.3 (18.4) | 67.6 (17.6) | 63.7 (18.8) | 66.7 (16.5) | 64.9 (18.9) |
| **Female sex \*\*** | 200 (48.0) | 82 (46.9) | 118 (48.8) | 36 (38.7) | 164 (50.6) |
| **Sample Processing Time (hours) \*** | 9.7 (13.4) | 7.5 (6.5) | 11.3 (16.5) | 7.0 (6.3) | 10.4 (14.8) |
| **Hospital Site \*\*** |  |  |  |  |  |
| CHUM | 105 (25.2) | 42 (24.0) | 63 (26.0) | 32 (34.4) | 73 (22.5) |
| JGH | 312 (74.8) | 133 (76.0) | 179 (74.0) | 61 (65.6) | 251 (77.5) |
| **Diabetes \*\*** |  |  |  |  |  |
| No | 288 (69.1) | 100 (57.1) | 188 (77.7) | 54 (58.1) | 234 (72.2) |
| Yes | 127 (30.5) | 75 (42.9) | 52 (21.5) | 39 (41.9) | 88 (27.2) |
| **Chronic Obstructive Pulmonary Disease \*\*** |  |  |  |  |  |
| No | 358 (85.9) | 145 (82.9) | 213 (88.0) | 77 (82.8) | 281 (86.7) |
| Yes | 57 (13.7) | 30 (17.1) | 27 (11.2) | 16 (17.2) | 41 (12.7) |
| **Chronic Kidney Disease \*\*** |  |  |  |  |  |
| No | 363 (87.1) | 151 (86.3) | 212 (87.6) | 77 (82.8) | 286 (88.3) |
| Yes | 52 (12.5) | 24 (13.7) | 28 (11.6) | 16 (17.2) | 36 (11.1) |
| **Congestive Heart Failure \*\*** |  |  |  |  |  |
| No | 355 (85.1) | 152 (86.9) | 203 (83.9) | 80 (86.0) | 275 (84.9) |
| Yes | 60 (14.4) | 23 (13.1) | 37 (15.3) | 13 (14.0) | 47 (14.5) |
| **Hypertension \*\*** |  |  |  |  |  |
| No | 179 (42.9) | 67 (38.3) | 112 (46.3) | 31 (33.3) | 148 (45.7) |
| Yes | 235 (56.4) | 107 (61.1) | 128 (52.9) | 61 (65.6) | 174 (53.7) |
| **Liver disease \*\*** |  |  |  |  |  |
| No | 403 (96.6) | 172 (98.3) | 231 (95.5) | 91 (97.8) | 312 (96.3) |
| Yes | 12 (2.9) | 3 (1.7) | 9 (3.7) | 2 (2.2) | 10 (3.1) |
| **Smoking Status \*\*** |  |  |  |  |  |
| Current Smoker | 13 (3.1) | 6 (3.4) | 7 (2.9) | 5 (5.4) | 8 (2.5) |
| Ex-smoker | 47 (11.3) | 21 (12.0) | 26 (10.7) | 11 (11.8) | 36 (11.1) |
| Never smoker | 233 (55.9) | 98 (56.0) | 135 (55.8) | 40 (43.0) | 193 (59.6) |
| **Dataset** | **Testing Cohort (Mount Sinai)** |  |  |  |  |
| COVID-19 Severity |  | Severe |  | Critical |  |
|  | All Samples<br>(n=569) | Cases<br>(n=392) | Controls<br>(n=177) | Cases<br>(n=233) | Controls<br>(n=336) |
| **Age in years \*** | 59.6 (19.4) | 63.0 (17.1) | 52.1 (22.1) | 63.7 (16.7) | 56.8 (20.6) |
| **Female sex \*\*** | 238 (41.8) | 154 (39.3) | 84 (47.5) | 87 (37.3) | 151 (44.9) |

\* Mean (SD)
\*\* N (%). When the counts of each cell do not sum to the total sample size, this is due to missing data or the patient answering, "I do not know".
Severe - Cases: individuals who tested positive for COVID-19 and died or required any type of respiratory support (including oxygen delivered by nasal prongs). Controls: individuals with COVID-19 but did not meet severe case criteria or individuals who presented with symptoms of COVID-19 but were SARS-CoV-2 PCR negative.
Critical - Cases: individuals who tested positive for COVID-19 and died or required respiratory support (intubation, continuous positive airway pressure, bilevel positive airway pressure, continuous external negative pressure, or high flow positive end expiratory pressure oxygen).
Controls: individuals with COVID-19 but did not meet critical case criteria or individuals who presented with symptoms of COVID-19 but were SARS-CoV-2 PCR negative.
Sample Processing Time: time in hours between sample collection and sample freezing.
CHUM: Samples recruited from the Centre Hospitalier de l'Université de Montréal
JGH: Samples recruited from the Jewish General Hospital

For the definition of adverse COVID-19 outcomes, we focused on two levels of severity: 1) severe COVID-19 was defined as individuals who died or required any form of oxygen supplementation; and 2) critical COVID-19, defined as individuals who died or experienced severe respiratory failure (requiring non-invasive ventilation, high flow oxygen therapy, intubation, or extracorporeal membrane oxygenation). Detailed definitions of these adverse outcomes are described in **Methods**. The overall study design is shown in **Figure 1**, which outlines the training and testing stages of the study. Consistent with recent successful large-scale genetic studies, we defined controls as all participants not meeting case criteria^1^.

**Figure 1.**
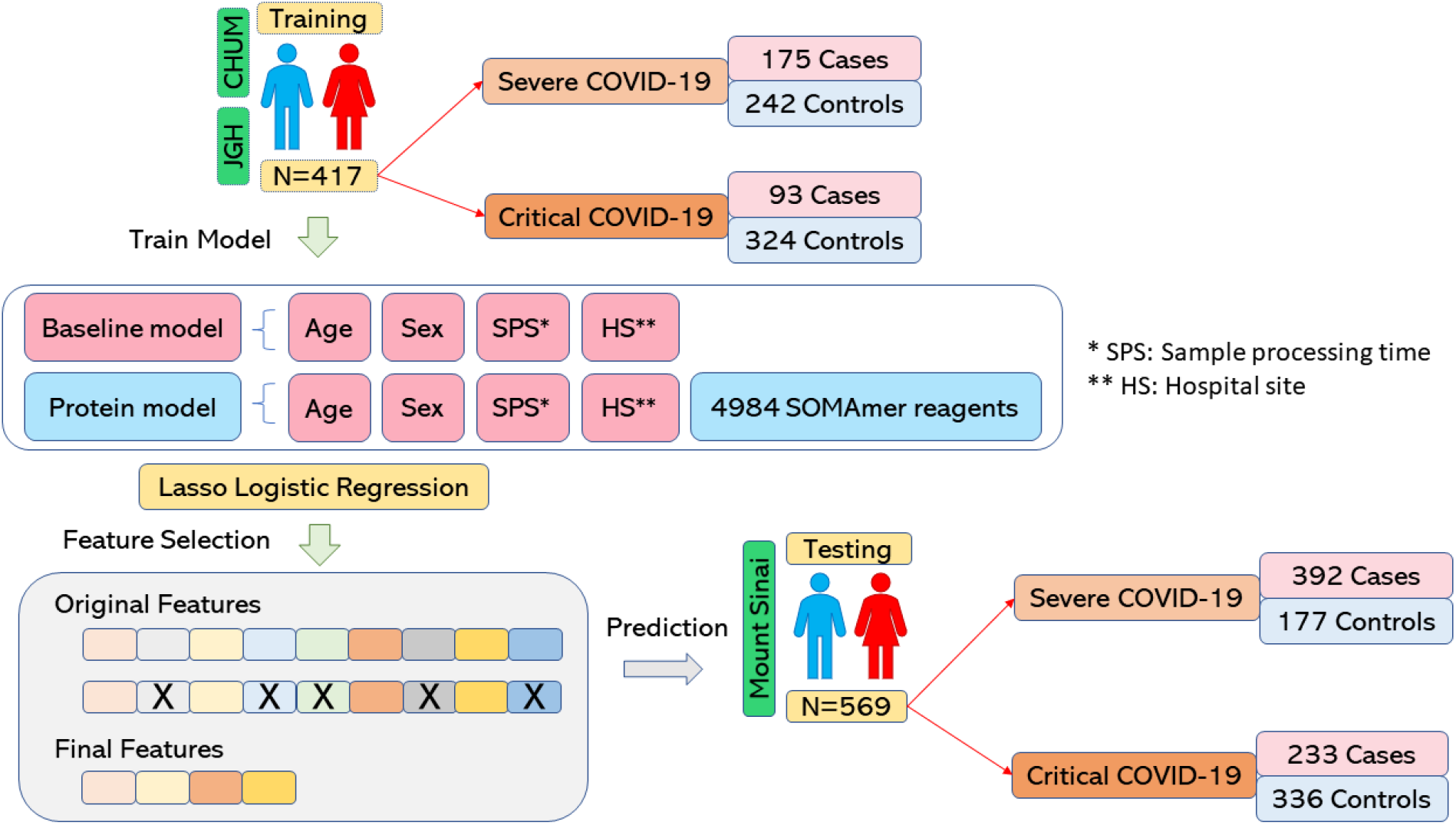
Overall Study design. Schematic of training and testing stages of this study. Severe COVID-19 is defined as death or use of any form of oxygen supplementation. Critical COVID-19 is defined as death or severe respiratory failure (non-invasive ventilation, high flow oxygen therapy, intubation, or extracorporeal membrane oxygenation).

In the BQC19 training cohort 175 individuals were classified as severe cases and 242 individuals were controls. The controls for severe COVID-19 were comprised of 138 SARS-CoV-2 positive individuals not meeting case definition and 104 SARS-CoV-2 negative individuals. In the case of critical disease, 93 individuals out of 313 COVID-19 positive patients were classified as critical cases and 324 individuals were controls. The controls for critical COVID-19 cases were 220 SARS-CoV-2 positive individuals not meeting case definition and 104 participants who were SARS-CoV-2 negative. In the Mount Sinai testing cohort, 392 individuals were classified as severe cases and 177 individuals were controls while for critical disease 233 individuals were cases and 336 were controls. Generally, severe, or critical COVID-19 cases were older than controls in both the training dataset and the testing dataset. Males were also more likely to have severe or critical COVID-19 as compared to females (**Table 1**). The age and sex distribution of the participants stratified by case/control status for the two COVID-19 severity outcomes are shown in **Supplementary Figure 1**. The distributions suggest that males who develop severe or critical COVID-19 are generally younger than females.

### Association of Protein Abundance with COVID-19 Outcomes

In order to directly assess if any of the measured proteins were associated with COVID-19 severity, we used multivariable logistic regression to test the association of each of the 4,984 SOMAmer reagents with the two COVID-19 outcomes while adjusting for age, sex, sample processing time, and hospital site in the BQC19 cohort. These variables were chosen because they are readily available in the course of clinical care, representing the minimum set of variables to predict outcomes. Logistic regression identified 1,531 SOMAmer reagents to be associated with severe COVID-19 (**Supplementary Table 1)** and 1,592 SOMAmer reagents (**Supplementary Table 2**) to be associated with critical COVID-19 when using a Benjamini-Hochberg corrected p-value of 0.01 (**Supplementary Figure 2**).

### Model Selection and Performance Using LASSO

One reason why many circulating proteins were associated with COVID-19 severity is that most of the protein levels were highly correlated with each other. Therefore, we used L1 regularized multivariable logistic regression models (LASSO)^40^ to select uncorrelated proteins that best predicted COVID-19 outcomes in the BQC19 training cohort. We did so for three reasons: 1) LASSO performs well when the number of features is greater than the number of samples (as was the case in our experiment); 2) LASSO forces many correlated features to have a zero coefficient by randomly selecting one of the features (sometimes more than one) from a group of correlated features thereby preventing collinearity; 3) LASSO mitigates the possibility of overfitting^40^.

We first defined a baseline model which included only the four covariates in the logistic regression model: age, sex, sample processing time, and hospital site to predict COVID-19 outcomes. We then evaluated whether the addition of proteins would aid in identifying which patients developed severe COVID-19 by adding 4984 SOMAmer reagents to the baseline model. This model, which included baseline covariates and protein levels, is termed the “protein model”. To train both the baseline and the protein models, we performed 10 repeats of stratified 5-fold cross-validation using LASSO logistic regression in the BQC19 cohort on both the severe and critical outcomes. We tuned the penalty parameter “lambda” across each of the 50 cross-validations and selected the lambda value corresponding to the model with the highest area under the receiver operator characteristic curve (AUC), which was averaged over the 50 cross-validation results. Results from the lambda parameter search are shown in **Supplementary Figure 3A-B**.

For the best performing model predicting the severe COVID-19 outcome, we selected a log_10_ lambda value of -1.5 which generated an average training AUC of 59% for the baseline model. We next selected a log_10_ lambda value of 1.0, which generated an average AUC of 88% for the protein model. For the best performing model predicting the critical COVID-19 outcome, we selected log_10_ lambda values of -2.0 and 1.0 corresponding to average cross-validation training AUC scores of 59% and 89% for the baseline and protein model, respectively (**Figure 2A-B**). We then used these chosen lambda hyperparameters to build baseline and protein models for severe and critical COVID-19 using the entire BQC19 cohort and evaluated their performance in the independent external test cohort from Mount Sinai.

**Figure 2.**
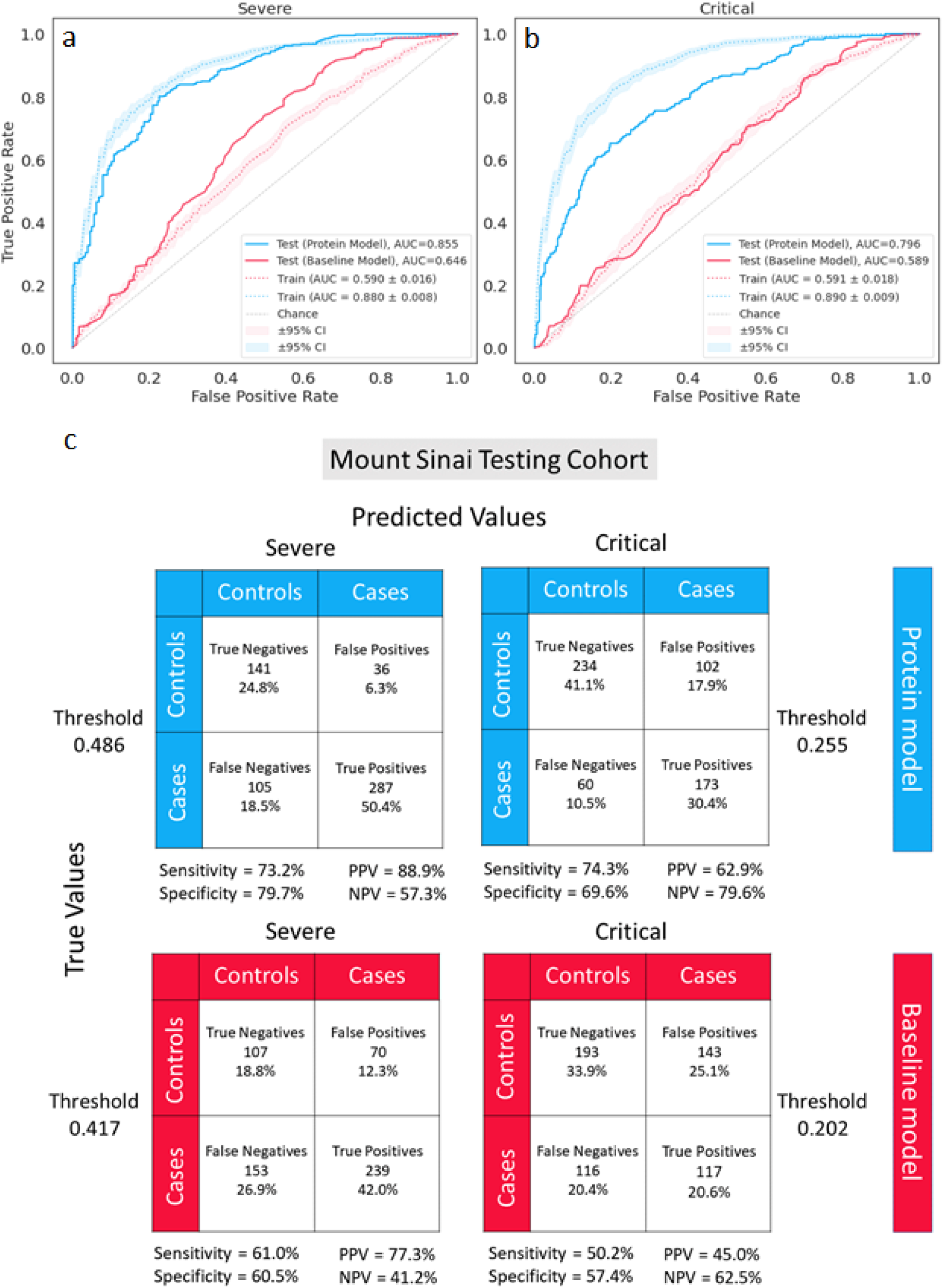
AUC score results. **a**, L1 regularized logistic regression training and testing results for severe COVID-19 and **b**, critical COVID-19. Blue and red are used to represent the protein model and baseline model, respectively, while solid and dotted lines represent the testing and training performance, respectively. Shaded areas denote the 95% confidence intervals for the training cohort. **c**, Two-by-two contingency table results from the test set are shown for predicting severe (top left, bottom left) and critical COVID-19 (top right, bottom right) using the protein model (blue) and baseline model (red). The threshold for predicting cases was determined during training using Youden’s J statistic which selects a threshold that maximizes the sum of the sensitivity and specificity score. PPV = positive predictive value, NPV = negative predictive value.

When testing the prediction of severe COVID-19 in the independent Mount Sinai cohort, AUC performance of the baseline model improved from 59% in the BQC19 training cohort to 65% in the Mount Sinai testing cohort. The AUC of the protein model decreased slightly between training and testing (88% vs. 86%).

The AUC of the protein model for predicting critical COVID-19 also decreased from a training score of 89% to 80% in the test set. In contrast, prediction of the critical COVID-19 outcome using the baseline model was consistent between training and test performance (AUC: 59%). The stability of these AUC estimates in the test cohort suggested that both protein models were robust.

The classification performance of the baseline and protein models in the Mount Sinai cohort is shown as two-by-two contingency tables in **Figure 2C**. The baseline and protein models used thresholds of 0.417 and 0.486 to predict severe COVID-19, respectively. These thresholds were selected by computing Youden’s J statistic during training and determine the threshold that maximized the sum of the sensitivity and specificity scores during training. The thresholds selected were roughly consistent with the case to control ratio in the BQC19 cohort used for training (175 cases, 242 controls). The protein model achieved a sensitivity of 73.2% compared to 61.0% for the baseline model, and a specificity of 79.7% compared to 60.5% for the baseline model, when predicting the severe COVID-19 outcome (**Figure 2C**).

When predicting critical COVID-19 using the baseline and protein models, thresholds of 0.202 and 0.255 were used to predict cases, respectively, using the same method. The low threshold for predicting critical COVID-19 cases is consistent with the case to control ratio in training which was 93 to 324 samples. The baseline model achieved a sensitivity / specificity score of 50.2% / 57.4% while the protein model achieved 74.3% / 69.6%, suggesting that the protein model trained to predict critical COVID-19 had decent power to classify true positives and true negatives (**Figure 2C**).

Furthermore, both the baseline and protein models demonstrated higher positive predictive values than negative predictive values when predicting the severe COVID-19 outcome, compared to the critical outcome. In contrast, these models produced higher negative predictive values than positive predictive values when predicting critical COVID-19.

Overall, these results suggest that the protein models predicting severe and critical COVID-19 both perform reasonably well in terms of the trade-off between sensitivity and specificity. The protein model is sensitive (73.2%) at identifying severe COVID-19 cases and similarly sensitive (74.3%) at identifying critical COVID-19 cases. Further, the positive predictive value for severe COVID-19 was high at 88.9%, while the negative predictive value was 57.3% (**Figure 2C**). These results suggest that a protein model could predict severe COVID-19 with relatively high confidence.

### Proteins Prioritized by LASSO to Predict COVID-19 Severity Outcomes

To predict the severe COVID-19 outcome, the best performing protein model selected 92 proteins along with age and sample processing time (**Figure 3, Supplementary Table 3**). Assessing the correlation of all 92 proteins, we found that, as expected, most of the proteins did not correlate with each other (mean absolute Spearman’s ρ = 0.17) **(Supplementary Figure 5A)**. Of 8464 total correlations (92 × 92), 8372 correlations (98.9%) had Spearman’s absolute ρ < 0.8.

**Figure 3.**
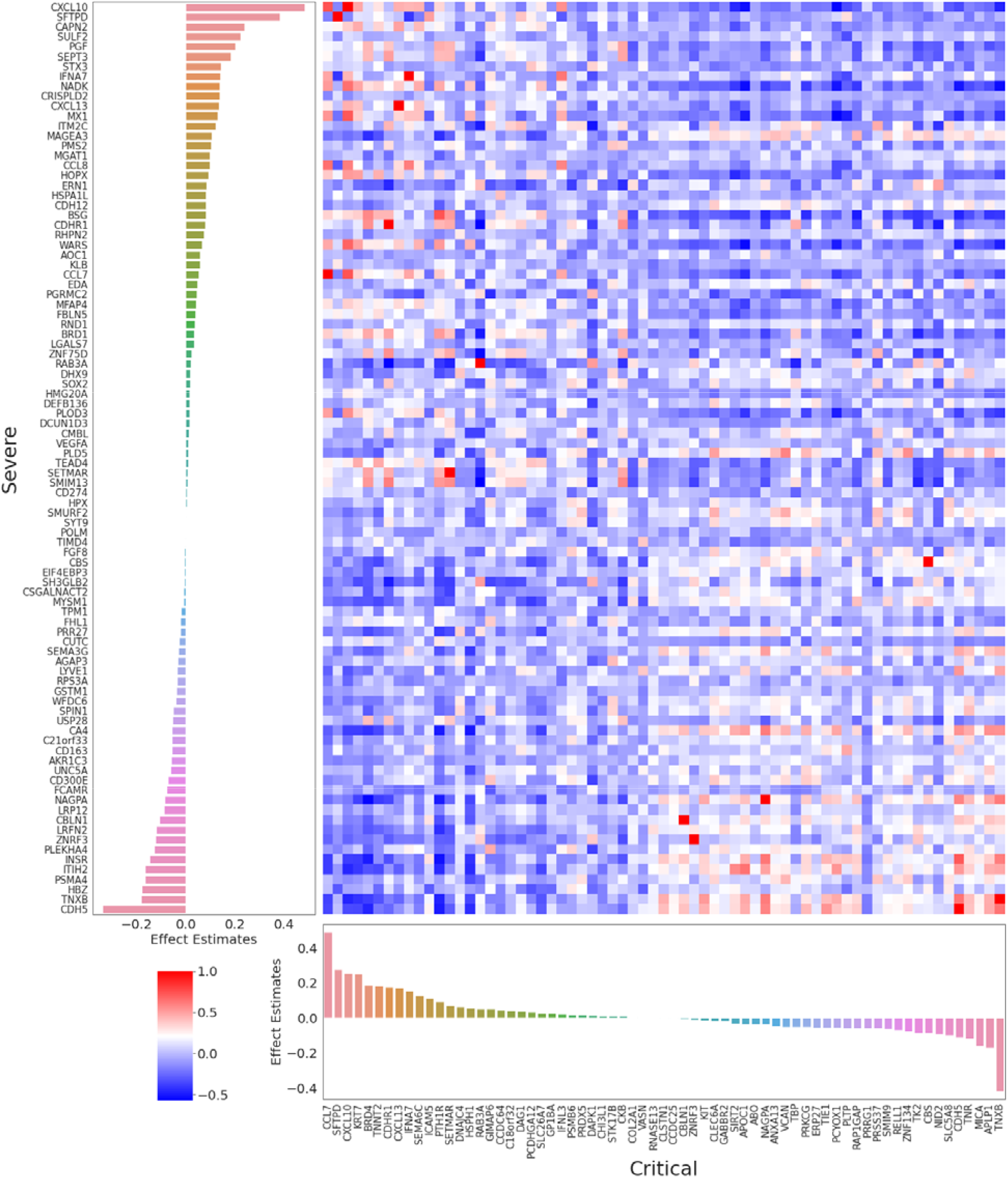
Feature importance and correlation of SOMAmers selected in the protein model to predict severe and critical COVID-19. (Left) Coefficient values of the 92 nonzero SOMAmer reagents in the final trained L1 regularized logistic regression protein model fitted on the severe COVID-19 outcome. The original data contained 4984 SOMAmer reagents and 4 other variables: age, sex, sample processing time, and hospital site. 92 SOMAmer reagents remained within the model along with age and sample processing time which are not shown. The model was trained on the entire BQC19 cohort using lambda = 10.0 (log_10_ lambda = 1.0) which was the best lambda value found from the hyperparameter search. (Bottom) Coefficient values of the 67 nonzero SOMAmer reagents of the final trained L1 regularized logistic regression protein model fitted on the critical COVID-19 outcome. The original data contained 4984 SOMAmer reagents and 4 variables age, sex, sample processing time, and hospital site. 67 SOMAmer reagents remained within the model along with age and sample processing time which are not shown. The model was trained on the entire BQC19 cohort using lambda = 10.0 (log_10_ lambda = 1.0) which was the best lambda value found from the hyperparameter search. (Right) Spearman’s rank correlations between the 92 proteins associated with the severe COVID-19 outcome and the 67 proteins associated with the critical COVID-19 outcome. These results suggest that while there were 14 overlapping proteins (SFTPD, CXCL10, RAB3A, NAGPA, CDH5, IFNA7, ZNRF3, CBS, CCL7, SETMAR, TNXB, CDHR1, CXCL13, and CBLN1), in general, the protein levels were uncorrelated with one another. Out of 6164 total correlations (92 × 67), 6150 correlations (99.8%) had a Spearman’s absolute ρ < 0.8.

Next, when predicting the critical COVID-19 outcome, the best performing protein model retained age, sample processing time, and 67 proteins (**Figure 3, Supplementary Table 4**). The absolute effect estimates of these proteins were generally larger than the severe COVID-19 model proteins (mean: 0.081 vs. 0.077). As expected, the 67 selected proteins also showed low levels of correlation (mean absolute Spearman’s ρ = 0.15) (**Supplementary Figure 5B)**. Of 4489 total correlations (67 × 67), 4422 correlations (98.5%) had Spearman’s absolute ρ < 0.8. The correlation between the 92 and 67 proteins selected to best predict severe and critical COVID-19 outcomes is shown in **Figure 3**. Out of 6164 total correlations (92 × 67), 6150 correlations (99.8%) had Spearman’s absolute ρ < 0.8. In general, proteins selected for predicting severe versus critical COVID-19 were not highly correlated (mean absolute Spearman’s ρ = 0.15). A hierarchically clustered heatmap after removal of the 14 common proteins in severe and critical COVID-19 showed that the proteins selected in predicting both outcomes were also generally uncorrelated with one another where 99.2% of the correlations had Spearman’s absolute ρ < 0.8 (**Supplementary Figure 5C**).

The percent of selected proteins for the prediction of severe and critical COVID-19 that were cytokines or chemokines was only 5.4% and 4.5%, respectively. Cytokine IFNA7, as well as chemokines CXCL13, CXCL10, CCL7, and CCL8 were present in the proteins selected for predicting severe COVID-19. Three chemokines, CXCL13, CXCL10, and CCL7 were selected for predicting critical COVID-19. Importantly, among the 14 overlapping proteins, those three chemokines, CXCL13, CXCL10, and CCL7 were selected for predicting both severe and critical COVID-19.

In addition, we clustered the 4,984 SOMAmer reagents using uniform manifold approximation and projection (UMAP) to reduce the feature space to a 2-dimensional space and highlighted the position of the model-selected proteins predictive of either severe or critical COVID-19. Our results suggested that LASSO selected proteins were sparsely distributed across the clusters **(Supplementary Figure 6**). This provides further evidence that: 1) few of the selected proteins are closely clustered with one another in UMAP space and 2) the proteins selected from the severe protein model and critical protein model were also quite distant from each other in UMAP space. Finally, we performed pathway analysis on common proteins with non-zero effect estimates that were included in the best predictive models of severe and critical COVID-19. Since LASSO is designed to pick one protein from a group of correlated proteins regardless of their biological relevance, we also included proteins highly correlated (Spearman’s absolute ρ > 0.75) with the proteins selected by LASSO for the enrichment analysis. As a result, 171 proteins were included in the severe group (92 LASSO selected proteins and 79 correlated proteins, **Supplementary Table 5**); and 96 proteins were included in the critical group (67 LASSO selected proteins and 29 correlated proteins, **Supplementary Table 6**). Among which, 32 proteins were common between the severe and critical groups.

We found that these 32 proteins were enriched in 35 pathways (g:SCS adjusted p value < 0.05), among which 15 were directly related to immune responses (**Supplementary Figure 7, Supplementary Table 7**). Prominent pathways included viral protein interaction with cytokines, cytokine and cytokine receptors (IL22RA1, TNFRSF10B, CCL7, CXCL10, CXCL13, adjusted P=0.0008) and cytokine-cytokine receptor interactions (CD4, IL22RA1, TNFRSF10B, CCL7, CXCL10, CXCL13, IFNA7, adjusted P= 0.002).

Interestingly, more than half of enriched pathways were not related to immune response (e.g., signaling receptor binding, cell activation). This suggests that other non-immune response pathways influence COVID-19 severity. In addition, some of these pathways included protein phosphorylation (CTSG, PRKCZ, PECAM1, CD4, CLEC7A, NPPA, TNFRSF10B, PRDX4, EPHA4, TNXB, CXCL10, IFNA7, CDH5) and glycosaminoglycan binding (CTSG, CCL7, TNXB, CXCL10, CXCL13), suggesting potential avenues to explore for drug development.

### Using Clinical Risk Factors to Predict COVID-19 Outcomes

In order to contrast the prediction capabilities of protein levels with established clinical risk factors, we performed two sensitivity analyses with results shown in **Table 2**. In the first analysis, we added six clinical risk factors to the baseline and protein models described in the main analyses. These clinical risk factors were diabetes, chronic obstructive pulmonary disease (COPD), chronic kidney disease, congestive heart failure, hypertension, and liver disease. The prevalence of these risk factors is shown in **Table 1**.

**Table 2.** Training AUC comparison.

|  | **Main Analysis**<br>**(n=417)** |  | **Main Analysis plus 6**<br>**clinical risk factors**<br>**(n=417)** |  | **Main Analysis plus 7**<br>**clinical risk factors**<br>**(n=312)** |  |
| --- | --- | --- | --- | --- | --- | --- |
|  | **Baseline**<br>**model** | **Protein**<br>**model** | **Baseline**<br>**model + 6**<br>**CRFs** | **Protein**<br>**model + 6**<br>**CRFs** | **Baseline**<br>**model + 7**<br>**CRFs** | **Protein**<br>**model + 7**<br>**CRFs** |
| **Severe COVID-19** | 0.590 | 0.880 | 0.636 | 0.880 | 0.657 | 0.848 |
| **Critical COVID-19** | 0.591 | 0.890 | 0.607 | 0.890 | 0.606 | 0.848 |

Addition of these six clinical features to the baseline model improved the training AUC to 64% (from 59%) when predicting severe COVID-19 and to 61% (from 59%) when predicting critical COVID-19. However, adding these clinical risk factors to the protein model resulted in no change in the training AUC performance when predicting severe COVID-19 (AUC = 88% vs. 88%) and critical COVID-19 (AUC = 89% vs. 89%) (**Table 2, Supplementary Figure 8**). 95 and 69 features with non-zero beta coefficient effect estimates were selected for the protein models predicting severe and critical COVID-19, respectively, in this sensitivity analysis (**Supplementary Figure 8**). Comparing proteins selected by the protein model in this sensitivity analysis, only one protein, KIT, was added to the 94 features selected in the main analysis. For critical COVID-19, the 69 features selected remained the same.

For the second sensitivity analysis, we augmented the first sensitivity analysis with an extra covariate for smoking status. Due to missing smoking information from the CHUM hospital site in the BQC19 cohort, only 312 samples were used in model training for the second sensitivity analysis. The results suggested that the addition of smoking and 6 clinical risk factors into the original baseline model composed of age, sex, sample processing time, and hospital site also slightly improved the training performance when predicting the severe COVID-19 outcome (AUC = 66% vs. 59%) and the critical COVID-19 outcome (AUC = 61% vs. 59%) (**Table 2, Supplementary Figure 9**). When adding smoking and these 6 clinical risk factors to the protein model, we found that training performance actually decreased for the severe COVID-19 outcome (AUC = 85% vs. 88%) and critical COVID-19 outcome (AUC = 85% vs. 89%). The non-zero beta coefficients of the proteins for severe and critical COVID-19 outcomes are shown in **Supplementary Figure 9** with a total of 79 and 51 features being selected, respectively. Comparing the 79 features selected by the protein model in this sensitivity analysis to the original 94 features selected previously when predicting severe COVID-19, we observed that only 48 features overlapped. Similarly, the 51 features selected by the protein model in this sensitivity analysis only had 28 features overlapping with the 69 features selected previously. The observed decrease in AUC and fewer number of overlapping proteins when comparing main analyses and sensitivity analyses may be due to the reduction in sample size used for training.

The results from these sensitivity analyses suggest that the protein measurements are likely able to act as partial proxies of the tested clinical risk factors. The addition of the clinical risk factors that we assessed may improve the predictive performance for both COVID-19 severity outcomes when only demographic and sample processing parameters are available. However, when protein measurements are available, adding these extra clinical risk factors may add little for improving predictions.

## Discussion

In this large-scale study testing the association of 4,701 circulating proteins with severe and critical COVID-19 outcomes, we found that a subset of these proteins were strong predictors of COVID-19 severity. Specifically, developing a model in 417 individuals and testing its performance in 569 separate samples from an independent external cohort, we demonstrated that a proteomic model was able to predict severe COVID-19, defined as requiring the use of oxygen, with an AUC of 86% and a positive predictive value of 89%. The addition of several commonly used clinical risk factors for COVID-19 severity did not improve the performance of this model. The identified proteins were strongly enriched for cytokine signaling and immune pathways, but also highlighted non-immune pathways. Taken together, these findings demonstrate that circulating protein abundances are able to predict COVID-19 outcomes with reasonable accuracy.

By including an independent cohort in this study, we implemented best practices for model development and validation^41^. An important aspect of any prediction model is the testing of the model in a cohort separate from the training cohort. Therefore, a strength of this study was that our samples were recruited from three separate hospitals, across two separate health care systems in two different countries. In this study, we used the same clinical risk factors and the exact same proteomic measurement procedure to both train and test the models. This increases the probability that the results presented are generalizable and not overfitted to the training data^42^. Indeed, for the severe COVID-19 outcome, there was little change in the AUC when comparing the training and test cohorts (88% vs. 86%).

Further, most studies that have tested the association between protein levels and COVID-19 outcomes have focused on circulating cytokines and chemokines^13,43–49^. While this is a reasonable approach given the nature of the disease, we are unaware of any other studies that have tested the association of 4,701 circulating proteins with COVID-19 outcomes. A recent study assessing thousands of proteins and their associations with COVID-19 outcomes achieved an AUC of 85%, but this was not tested in an independent cohort^30^.

Interestingly only 5 of the 14 proteins selected in the final model of both severe and critical COVID-19 outcome were cytokines or chemokines. There were also proteins selected that were not specific to immune pathway proteins, such as glycosaminoglycan binding—a favourable set of targets for drug development. This suggests that many of the biological pathways that influence severity of COVID-19 outcomes may act distinctly from known cytokine and chemokine proteins.

A major clinical challenge within the pandemic has been the triaging of patients to identify those most likely to require admission for hospitalization^50^. A common reason for hospitalization is the need for oxygen support. Currently, treating physicians are required to assess the need for admission using models with poor predictive performance. A model generated in China early in the pandemic to predict COVID-19 severity requires a medical history, chest X-ray, and extensive blood testing^51^. Further, the 4C Mortality Score was able to predict in-hospital mortality, but achieved an AUC of only 77%^52^. Approximately half of the patients enrolled in our study have developed severe or critical COVID-19 after their baseline proteomic measurement which were used as predictors in this study. This suggests circulating protein measurements could be considered for predicting COVID-19 severity, but this requires further study, including more sampling at the onset of symptoms.

This study has important limitations. While the model was tested in a separate cohort, and generalized well, it should be tested in additional cohorts, especially in cohorts of diverse ancestry. The control population included individuals who were SARS-CoV-2 positive and had mild disease, in addition to individuals who were suspected to have COVID-19 but were SARS-CoV-2 negative. This means that the developed models provide insight into prediction of individuals who develop severe COVID-19 compared to mild COVID-19 and other acute diseases having symptoms consistent with COVID-19. Such control definitions reduce the potential for collider bias, but do not allow direct prediction of COVID-19 outcomes amongst only COVID-19 patients^53^. Last, the clinical translation of this study is hindered by the cost involved in measuring 4,701 circulating proteins but could be improved by developing a specific assay to the selected proteins.

In summary, circulating protein levels are strongly associated with COVID-19 outcomes and able to predict the need for oxygen supplementation or death with reasonable accuracy. Measured protein levels were superior to predicting COVID-19 severity outcomes when compared to nearly all clinical risk factors tested. Further research is needed to assess whether this proteomic approach can be applied in a clinical setting to assist in triaging patients for admission to hospital.

## Data Availability

Code used in this analysis is available at https://github.com/chenyangsu/somalogic with additional information available upon request. The BQC19 is an Open Science Biobank. Instructions on how to access data for individuals from the BQC19 at the Jewish General Hospital site is available at https://www.mcgill.ca/genepi/mcg-covid-19-biobank. Instructions on how to access data from other sites of the BQC19 is available at https://www.bqc19.ca/en/access-data-samples.

## Ethics declarations

All contributing cohorts to the present analyses received ethics approval from their respective research ethics review boards. The Biobanque Québécoise de la COVID-19 (BQC19) received ethical approval from the IRB of the JGH and the CHUM. This research was reviewed and approved by the Icahn School of Medicine at Mount Sinai Program for the Protection of Human Subjects (PPHS) under study number 20-00341. This research was considered minimal risk Human Subjects Research.

## Author contributions

Conception and design: CYS, SZ, WJ, JP, and JBR. Data analyses: CYS, SZ, and EGK. Data acquisition: TN, GBL, DM, DEK, JA, MA, LL, EBR, DH, NK, ZA, NR, MB, LP, CG, XX, CT, BV, OA, TA, NA, NB, MD, KN, NWS, KM, DMDV, NZ, MP, HX, JH, RM, EC, KT, KA, IS, NB, EK, VF, TM, SG, SKS, AC, MM, DEK, and JBR. Interpretation of data: CYS, SZ, EGK, GBL, EBR, TN, DEK, and JBR. Funding acquisition: VM, TM, SG, SKS, AC, MM, DEK, and JBR. Methodology: CYS, SZ, EGK, CMTG, CP, MH, JCZS, CL, and JBR. Project administration: DM, VF, AC, MM, DEK, JBR. Validation: CYS, SZ, GBL, EGK, EBR, YC, YF, DEK, and JBR. Visualization: CYS and SZ. Writing-original draft: CYS, SZ, EGK, GBL, EBR, TN, DEK, and JBR. Writing-review & editing: CYS, SZ, EGK, GBL, EBR, TN, DEK, ES, MD, CP, VM, DEK, and JBR. All authors were involved in further drafts of the manuscript and revised it critically for content. All authors gave final approval of the version to be published. The corresponding author attests that all listed authors meet authorship criteria and that no others meeting the criteria have been omitted.

## Acknowledgements

The Richards research group is supported by the Canadian Institutes of Health Research (CIHR: 365825; 409511, 100558), the Lady Davis Institute of the Jewish General Hospital, the Jewish General Hospital Foundation, the Canadian Foundation for Innovation, the NIH Foundation, Cancer Research UK, Genome Québec, the Public Health Agency of Canada, McGill University, Cancer Research UK [grant number C18281/A29019], and the Fonds de Recherche Québec Santé (FRQS). The Kaufmann lab’s COVID-19 work is supported by the Canadian Institutes of Health Research /CITF (VR2-173203 and VS1-175561), the American Foundation for AIDS Research (AmFAR 110068-68-RGCV), the Canadian Foundation for Innovation, and FRQS. Support from Calcul Québec and Compute Canada is acknowledged. TwinsUK is funded by the Welcome Trust, Medical Research Council, European Union, the National Institute for Health Research (NIHR)-funded BioResource, Clinical Research Facility, and Biomedical Research Centre based at Guy’s and St Thomas’ NHS Foundation Trust in partnership with King’s College London. These funding agencies had no role in the design, implementation, or interpretation of this study. The measurement of proteomics using the SomaLogic panel was supported by the McGill Interdisciplinary Initiative in Infection and Immunity (MI4) and by the Bill and Melinda Gates Foundation for the Mount Sinai cohort.

J.B.R. and D.E.K. are supported by FRQS Mérite Clinical Research Scholarships. C.-Y.S. is supported by a Lady Davis Institute / TD Bank Studentship Award. S.Z. is supported by a CIHR fellowship and an FRQS postdoctoral scholarship. G.B.L. is supported by a CIHR scholarship and a joint FRQS and Québec Ministry of Health and Social Services scholarship. T.N. is supported by Research Fellowships of the Japan Society for the Promotion of Science (JSPS) for Young Scientists. M.D. is supported by a clinician-researcher salary award from the FRQS. V.M. is supported by a Canada Excellence Research Chair.

## Members of the Mount Sinai COVID-19 Biobank Team are listed in Supplementary Table 8

### Methods

#### Cohorts

The Biobanque Québécoise de la COVID-19 (BQC19) is a Québec-wide biobank which was launched to enable research into the causes and consequences of COVID-19 disease (see bqc19.ca)^54^. For this study, we used results from 417 patients (313 SARS-CoV-2 nasal swab PCR positive patients and 104 individuals who presented with symptoms consistent with COVID-19 but had negative SARS-CoV-2 PCR nasal swabs) with available proteomic data from the SomaScan SomaLogic® assay. The subjects were recruited at the Jewish General Hospital (JGH) and Centre Hospitalier de l’Université de Montréal (CHUM) in Montréal, Québec, Canada, both of which are university affiliated hospitals. For each individual, blood samples drawn at the earliest time point were used for training when an individual had multiple blood draws. Selecting the blood sample at the earliest time point reflects the protein measurements during the acute phase of COVID-19 disease. The demographic characteristics of the participants in the BQC19 cohort who underwent SomaScan® assays is detailed in **Table 1**. The demographic characteristics were obtained by medical chart review or patient interview performed by trained clinicians or trained research coordinators.

The Mount Sinai cohort used in this study was composed of results from 569 patients made up of 472 SARS-CoV-2 positive patients and 89 SARS-CoV-2 negative patients confirmed through PCR tests, one COVID-19 positive patient diagnosed by a chest CT while the remaining 7 individuals were COVID-19 negative and did not have COVID-19 symptoms during specimen collection but may have had a history of exposure. The samples donated by the patients in the Mount Sinai cohort underwent the same proteomic data collection and profiling performed as in the BQC19 cohort. The subjects were recruited at the Mount Sinai Hospital in New York City which is affiliated with the Icahn School of Medicine. **Table 1** lists the demographic and sample processing parameters of participants in the Mount Sinai cohort that underwent SomaScan® assays. Demographic characteristics were obtained similarly to that of the BQC19 cohort.

#### Demographic, Sample Processing, and Clinical Variable Definitions

Age and sex from the BQC19 and Mount Sinai cohorts were collected. Sample processing time and hospital site were collected for BQC19 samples with the former being a continuous variable that quantifies the time in hours between sample collection and sample freezing.

The clinical variables were collected for the BQC19 cohort only. Clinical variables included smoking status and six different comorbidities: diabetes, COPD, chronic kidney disease, congestive heart failure, hypertension, and liver disease. All seven variables were collected as categorical values with the six comorbidities having three options (0 No, 1 Yes, and -1 Don’t know) while smoking status contained 4 categories (0 Current Smoker, 1 Ex-smoker, 2 Never smoked, and -1 Don’t know).

#### Proteomic Measurement using the Somascan Platform

Blood samples from both the BQC19 and Mount Sinai cohorts were collected using acid citrate dextrose (ACD) tubes. Proteomic measurement was performed at Somalogic using the Somascan v4.0 platform. In the BQC19 cohort, a total 1,038 samples collected at different time points from 503 individuals were sent to SomaLogic for proteomic profiling as previously described^2^, while the Mount Sinai cohort contained 1200 samples collected at different time points from 592 individuals that were sent to SomaLogic for proteomic profiling.

SomaLogic uses the Somascan proteomic platform which provides measurements on 4,701 unique human circulating proteins using 4,987 Slow Off-Rate Modified Aptamers (SOMAmer reagents) and quantifies protein levels in the form of relative fluorescence units (RFUs). Normalization and calibration steps were performed by SomaLogic to remove any systematic biases stemming from raw assays or samples. The normalization procedure involved three steps performed in a non-consecutive fashion: hybridization control normalization, intraplate median signal normalization, as well as plate scaling and calibration. More details on SomaLogic normalization can be found in their Technical Note^55^.

#### Data Preprocessing

A per-sample normalization process involved using a scale factor for a set of SOMAmer reagents to compute against a reference value generated from the median of all calibrated, unnormalized samples, and then aggregating the results within a dilution. This was done because using a normal population reference generated from EDTA plasma tubes would have been inappropriate for normalization, since samples in this study were from ACD plasma tubes. Due to the nature of the samples that were collected from patients during acute infection, we did not apply the recommended scale [0.4-2.5] to remove samples. The raw dataset composed of 5284 SOMAmer reagents was first processed with SomaLogic package SomaDataIO v3.1.0. We removed any SOMAmer reagents that represented non-human proteins or controls (NoneX, NonHuman, Spuriomer, HybControlElution, NonBiotin, NonCleavable) and retained 4984 unique SOMAmer reagents for analysis.

#### Curation of Samples from the Longitudinal Dataset

To investigate our primary study question, we focused on samples collected during the acute infection stage. Samples from the acute infection stage were defined as samples collected from SARS-CoV-2 PCR positive patients within 14 days of symptom onset. When an individual provided multiple samples collected within 14 days of symptom onset, the sample collected at the earliest timepoint was retained for analyses. Both the BQC19 and Mount Sinai samples adhered to this rule.

#### COVID-19 Outcome Definitions

We defined two sets of severity outcomes for COVID-19: severe COVID-19 and critical COVID-19. Positive SARS-CoV-2 results were confirmed by SARS-CoV-2 viral nucleic acid amplification tests (NAAT) from relevant biologic fluids. Cases for severe COVID-19 were defined as individuals who tested positive for COVID-19 and died or required any type of respiratory support (including oxygen delivered by nasal prongs) at any timepoint. Controls for severe COVID-19 were defined as individuals who did not meet these severe case criteria; thus, controls were individuals with COVID-19 but did not meet severe case criteria or were individuals who presented with symptoms of COVID-19 but were SARS-CoV-2 PCR negative. Cases for critical COVID-19 were defined as individuals who tested positive for COVID-19 and died or required invasive respiratory support (intubation, continuous positive airway pressure, bilevel positive airway pressure, continuous external negative pressure, or high flow positive end expiratory pressure oxygen) at any timepoint. Controls for critical COVID-19 were individuals with COVID-19 but did not meet critical case criteria or were individuals who presented with symptoms of COVID-19 but were SARS-CoV-2 PCR negative.

#### Multivariable Logistic Regression

Multivariable logistic regression models were used to test the associations of either severe or critical COVID-19 on four covariates along with each SOMAmer reagent: age, sex, sample processing time, and hospital site. We used R package “glm” to perform 4984 logistic regression models in the BQC19 cohort. We first applied a false discovery rate of P < 0.01 (corrected P values were determined using the Benjamini-Hochberg procedure^56^, p.adjust with method set to “BH” in R) to select a subset of proteins associated with severe or critical COVID-19 outcomes. Volcano plots measuring the uncorrected -log_10_ P values as a function of the effect size estimates of each SOMAmer reagent were generated using the bioinfokit version 2.0.4 package in Python 3.7.

#### Regularized Logistic Regression Models

We defined two model types differing in the covariates used to train the model. The first model type is a “baseline model” which was trained using age, sex, sample processing time, and hospital site. The second model type is a “protein model” which is trained using age, sex, sample processing time, hospital site, and 4984 SOMAmer reagents. We used the baseline model in our analyses as a performance benchmark to compare the results of the protein model which we expected to perform better.

To predict the two COVID-19 severity outcomes defined above, we used LASSO regression and elastic nets. Specifically, for LASSO regression we used L1 Regularized Logistic Regression (Sparse Logistic Regression) as implemented in the “LogisticRegression” module from Sci-kit learn version 0.24.1, a machine learning library, with the penalty set to “L1”. The L1 norm penalty adds a constraint to the effect estimates of the regression model by setting many variables to have a null effect or a coefficient of 0. This in turn allows a form of feature selection to occur and also prevents overfitting to the training dataset by forcing the model to be less complex. In addition, when multiple variables are correlated with one another such as in the case of highly correlated proteins, the penalty term from LASSO may select a single variable from the group, thus allowing a subset of uncorrelated proteins to be selected. It is important to note that although LASSO tends to select a single variable from a group of highly correlated variables, this property is not a certainty. LASSO may occasionally select more than one variable depending on the size of the dataset and the value of the penalty term. To train this model, the hyperparameter “lambda”, which controls the amount of L1 regularization to add to the model, was first tuned through cross-validation (details are described in the next section). A larger value of lambda increases the amount of L1 regularization and forces more of the variables to have a null effect. On the other hand, training the model on a smaller lambda value will result in a model with more nonzero coefficients.

#### Cross-Validation and Hyperparameter Tuning

Due to the relatively small size of our training dataset from BQC19, we used 10 repeats of five-fold cross-validation to tune the hyperparameter, lambda, over 17 different lambdas (log_10_ values of lambda from -2 to 2, incremented by 0.25). Each repeat of the five-fold cross-validation process involved splitting the dataset into five folds: training on four folds and validating the trained model on the final fold and performing the process five times to cover each validation fold. We used a stratified cross-validation approach, meaning that the train and validation folds maintained the same percentage of samples of each class (case/control) as the original data. This is important because of the unbalanced case/control samples for the critical COVID-19 outcome (93 cases / 324 controls). A standard five-fold cross-validation split may result in train and validation folds with varying proportions of cases and controls. Since classification algorithms tend to weight each sample equally, the class that is overrepresented, such as the controls in the critical COVID-19 outcome, will receive more weight and thus bias the results. The stratified cross-validation step was performed using the RepeatedStratifiedKFold function in Sci-kit learn. Due to the relatively small sample size of our training set (*n*=417), we performed 10 repeats of this cross-validation process to stabilize the results from training. Each repeat first shuffled the entire training set then split the data into five folds which created more variability in the data used for training.

During training on the four folds, we standardized only the protein levels using the population standard deviation (i.e., dividing by the number of samples n) and use this mean and standard deviation to standardize the protein levels in the validation fold prior to validating. This prevents information leakage which can occur if standardization of protein levels was performed on the entire dataset rather than just the training folds. Age and sample processing time were treated as continuous variables, whereas sex and hospital site were treated using dummy variables (sex [0: Female, 1: Male], and hospital site [0: CHUM, 1: JGH]).

We used the AUC to determine the model performance during cross-validation. To select the best value for the hyperparameter lambda, we compared the average AUC score (computed from 50 validation fold results) for all lambda values and selected the lambda value corresponding to the highest average AUC. Youden’s J statistic was calculated for each receiver operator characteristic (ROC) curve during training. This performance metric can be calculated by subtracting the false positive rate from the true positive rate for each data point on a ROC curve and taking the maximum value. The threshold which corresponds to this maximum Youden’s J statistic is the threshold that maximizes the sum of the sensitivity and specificity for that particular ROC curve. We computed the threshold corresponding to the maximum Youden’s J statistic for each of the 50 ROC curves and averaged the 50 thresholds to get a single threshold value. This averaged threshold value was computed for each of the baseline and protein models predicting severe and critical COVID-19 and used to produce two-by-two contingency tables and therefore sensitivity and specificity values in the Mount Sinai cohort during model testing.

#### Model Testing

We checked the generalizability of the baseline and protein models by testing them in an external, independent dataset from Mount Sinai. Protein measurements in the test dataset were first natural log transformed then standardized using the mean and standard deviation of the corresponding protein in the training set. Similarly, age was not standardized and kept in years. Since samples were only from a single hospital, the hospital site parameter was left as is and did not need to be dummy encoded. The variable sample processing time, however, was absent from the testing set. For this reason, we imputed the sample processing time variable in the test cohort using the mean value of the sample processing time variable in the BQC19 training cohort.

#### Protein Correlations

Spearman’s Rank Order Correlation was used to determine the correlations between individual proteins in the BQC19 cohort. Heatmaps show magnitudes of correlation coefficients between values of -1 and 1. Correlation heatmaps showing collected clusters such as in **Supplementary Figure 5** were generated using the ggcorrplot function in R with the parameter hc.order set to TRUE to perform hierarchical clustering. Moreover, we reduced the dimension of the correlation matrix of 4984 SOMAmer reagents to a 2-dimensional space using uniform manifold approximation and projection (UMAP) from the umap-learn 0.5.1 package using default parameters. We annotated the SOMAmer reagents selected from the protein model that were associated with severe COVID-19 and critical COVID-19 as well as the proteins that overlapped between the two outcomes.

#### Pathway Enrichment Analyses

We used the web-based tool g:Profiler (https://biit.cs.ut.ee/gprofiler/gost) to investigate the possible pathways of the selected proteins as good predictors for both critical and severe COVID-19 identified by LASSO. The g:SCS algorithm was used to estimate the threshold for enrichment against all annotated genes. We selected pathways and interaction databases including Gene Ontology, KEGG, Reactome; TRANSFAC, miRTarBase, Human Protein Atlas, and CORUM.

#### Sensitivity Analyses

We tested the effect of six established clinical risk factors which included: diabetes, COPD, chronic kidney disease, congestive heart failure, hypertension, and liver disease in the BQC19 cohort to determine whether addition of comorbidities could improve prediction of COVID-19 severity outcomes. We added these six additional covariates, with characteristics shown in **Table 1**, to the baseline and protein models to perform LASSO regression analysis. A total of 417 samples from the BQC19 cohort were used for training.

We performed a second sensitivity analysis by adding smoking status along with these six established clinical variables to the baseline and protein models for LASSO regression analyses. Therefore, the baseline model contained covariates age, sex, sample processing time, hospital site, and seven clinical variables while the protein model contained all the baseline variables along with 4984 SOMAmer reagents. Since smoking status was not available from the CHUM hospital site, this sensitivity analysis only involved 312 samples from the BQC19 cohort that were collected at the JGH site.

Due to missing data, we imputed the values of samples: we first converted all six comorbidity features to binary values. Any value other than a “Yes” was converted to a “No” which may include missing values being converted to a “No”. For smoking status, we grouped all values into three categories: 0 - Current Smoker, 1 - Ex-smoker, and anything else (including missing values and -1) was set as 2 - Never smoked. Smoking status was dummy encoded and had one of the encoded variables dropped to prevent collinearity. For both sensitivity analyses, training of the L1 regularized logistic regression models used 10 repeats of stratified five-fold cross-validation as in the primary analysis.

