## Supplementary Figures and Text for "Circulating proteins to predict adverse COVID-19 outcomes"

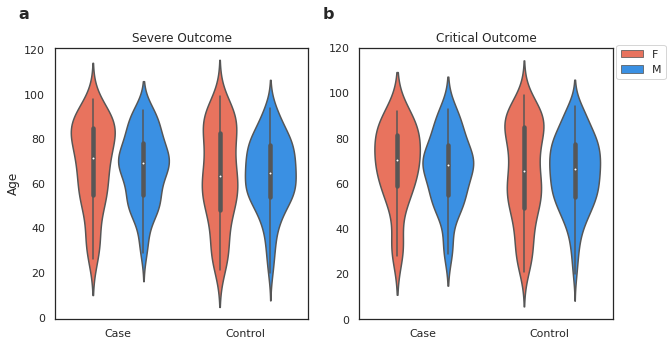

**Supplementary Figure 1** **|** **Violin plots showing the age and sex distribution of the BQC19 training cohort for severe and critical COVID-19.** **a**, Severe COVID-19. **b**, Critical COVID-19. Coloring indicates the sex of the patient as shown in the legend (F: Female, M: Male).

2a	2b

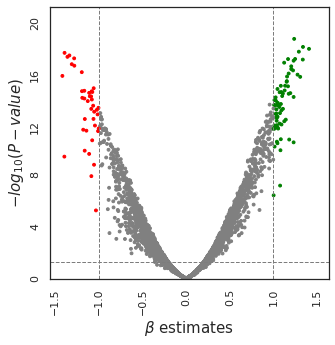

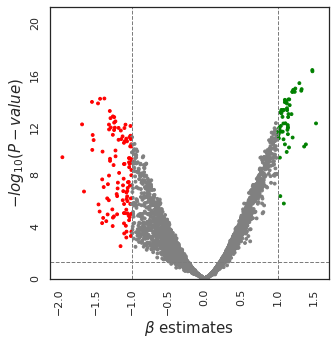

**Supplementary Figure 2** **|** **Volcano plots.** **a**, Severe COVID-19. **b**, Critical COVID-19. Each data point represents the association of a single SOMAmer reagent from multivariable logistic with these COVID-19 outcomes adjusted for age, sex, sample processing time, and hospital site. Vertical gridlines are set at beta effect estimates of -1.0 and 1.0 while the horizontal gridline represents the p value with a significant threshold set at P = 0.05. The beta effect estimate shows the change in the log odds of having severe or critical COVID-19 disease associated with a single standard deviation change in the SOMAmer reagent of interest. Statistically significant SOMAmer reagents with large beta estimate changes in the negative direction and positive direction are shown in red and green, respectively.
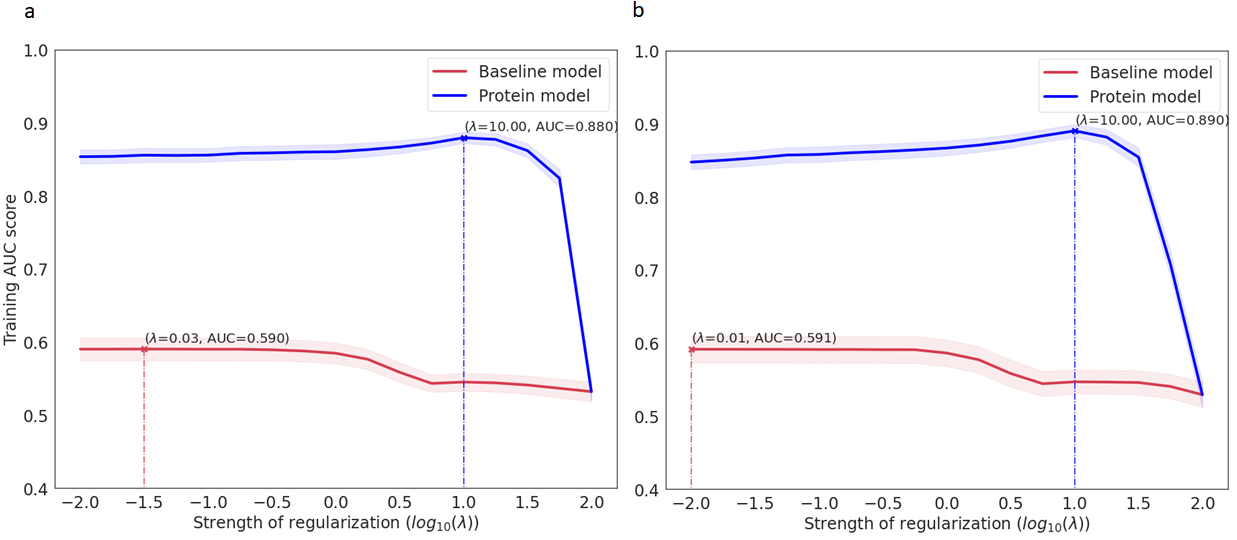

**Supplementary Figure 3 | AUC score as a function of regularization strength during L1 regularized logistic regression training to determine the best lambda hyperparameter value. a**, Severe COVID-19 outcome. **b**, Critical COVID-19 outcome. The baseline model (red) was fitted with covariates age, sex, sample processing time, and hospital site. The protein model (blue) was fitted with age, sex, sample processing time, hospital site, and 4984 SOMAmer reagents. Each data point represents an AUC score averaged over 50 validation folds. Shaded areas represent 95% confidence intervals above and below the mean AUC.

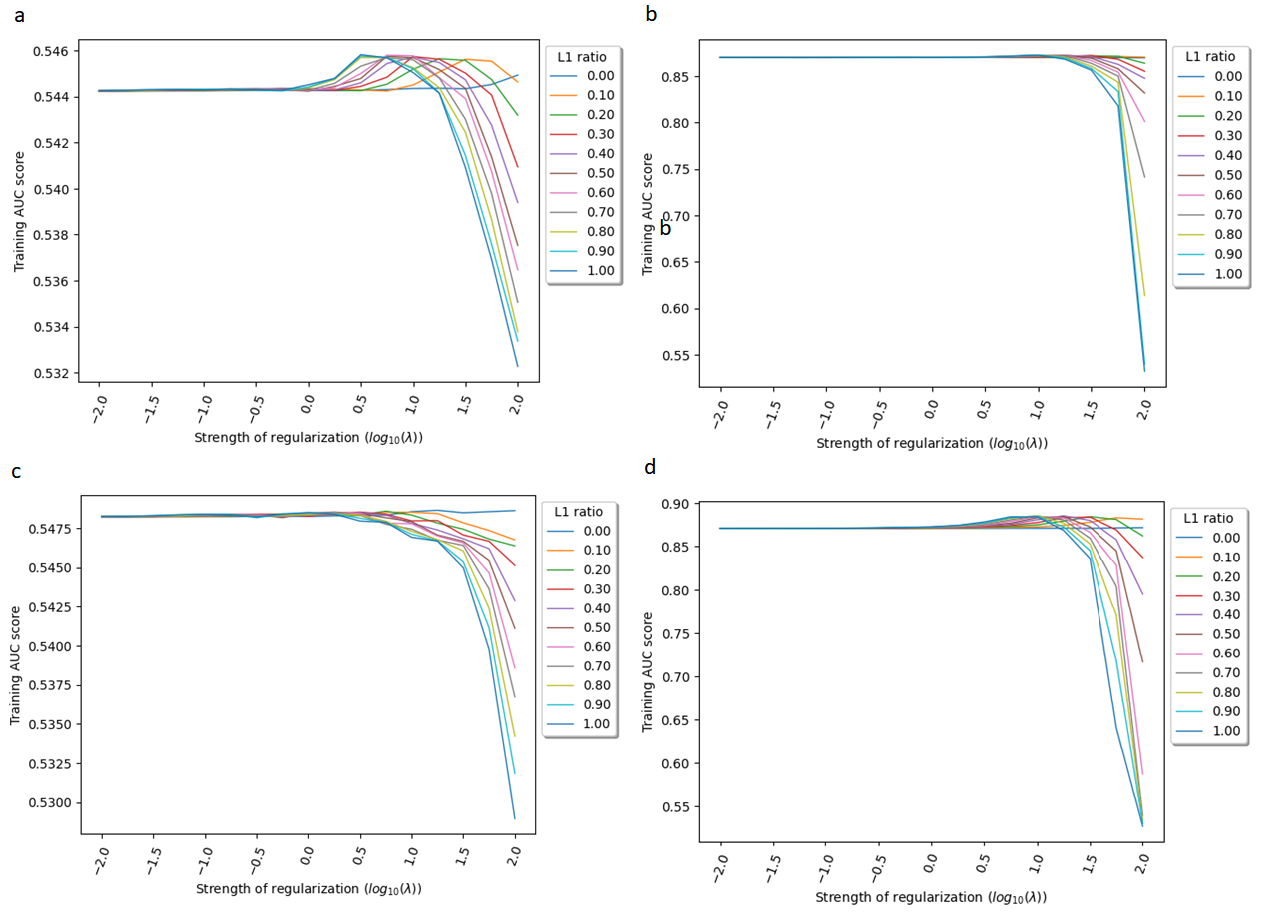

**Supplementary Figure 4 | Elastic net logistic regression hyperparameter search results.** AUC score as a function of regularization strength during elastic net regularized logistic regression training on severe COVID-19 for the **a**, baseline model and **b,** protein model. **c,** AUC score as a function of regularization strength during elastic net regularized logistic regression training on critical COVID-19 for the **c**, baseline model and **d,** protein model.

L1 ratio = 0 represents using L2 regularization fully i.e., Ridge Logistic Regression while L1 ratio = 1 represents using LASSO Logistic Regression. Each data point represents an AUC score averaged over 50 validation folds. Parameters to tune were the lambda value (same as in L1 regularized logistic regression) and the L1 ratio which represents a linear combination of L1 and L2 regularizations. Lambda is shown on the x-axis while the L1 ratio is shown as different colored lines. Training AUC is shown on the y-axis.

a
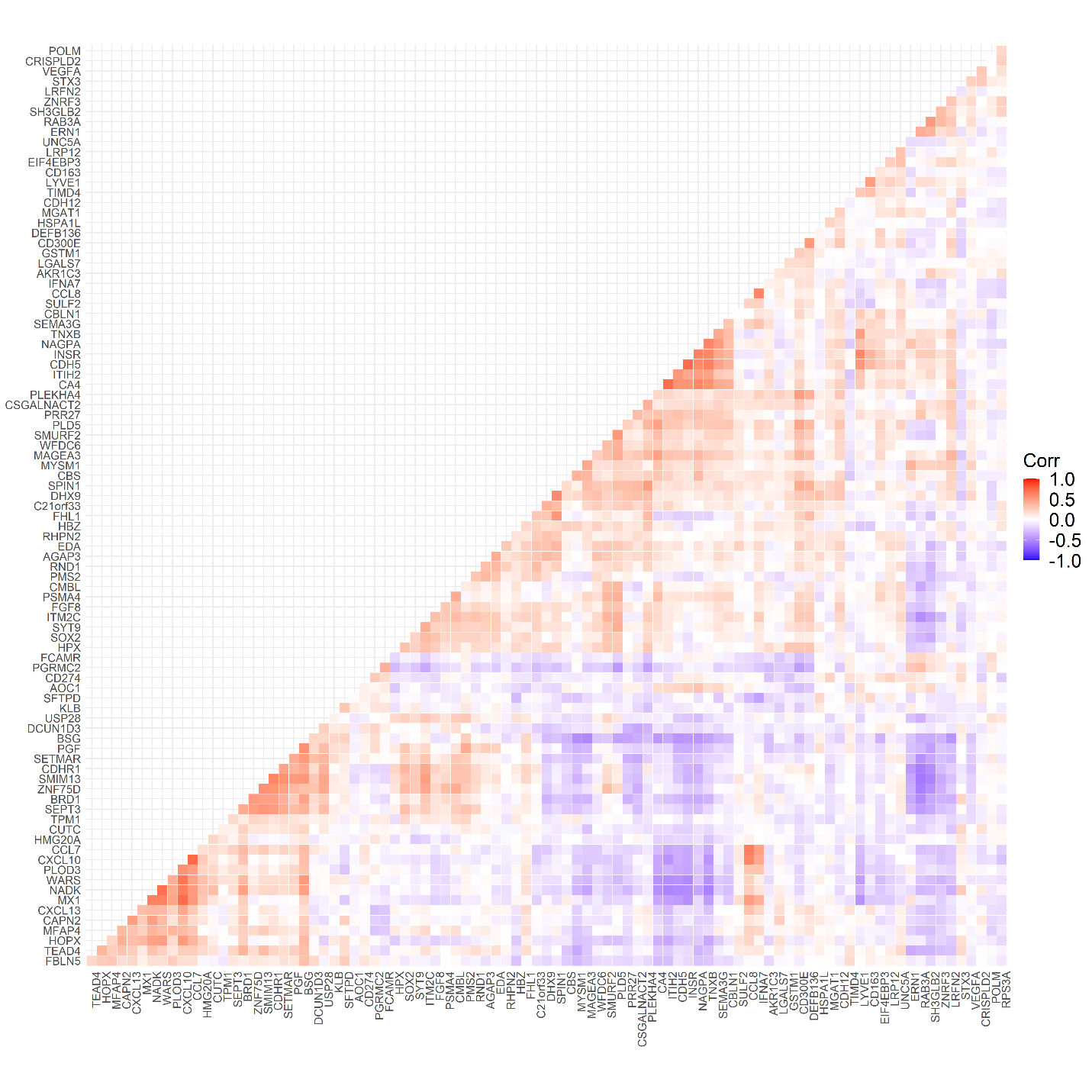

b
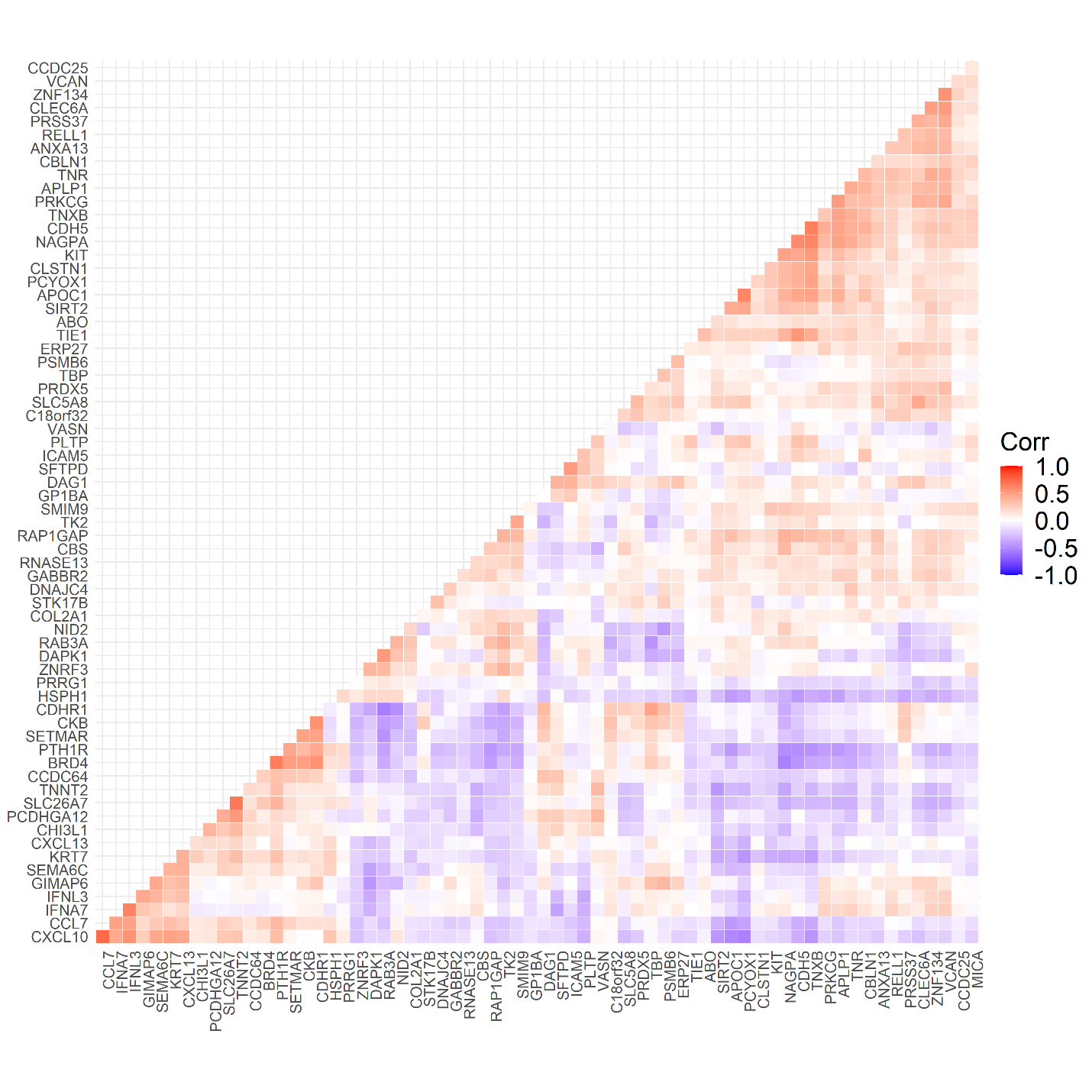

c
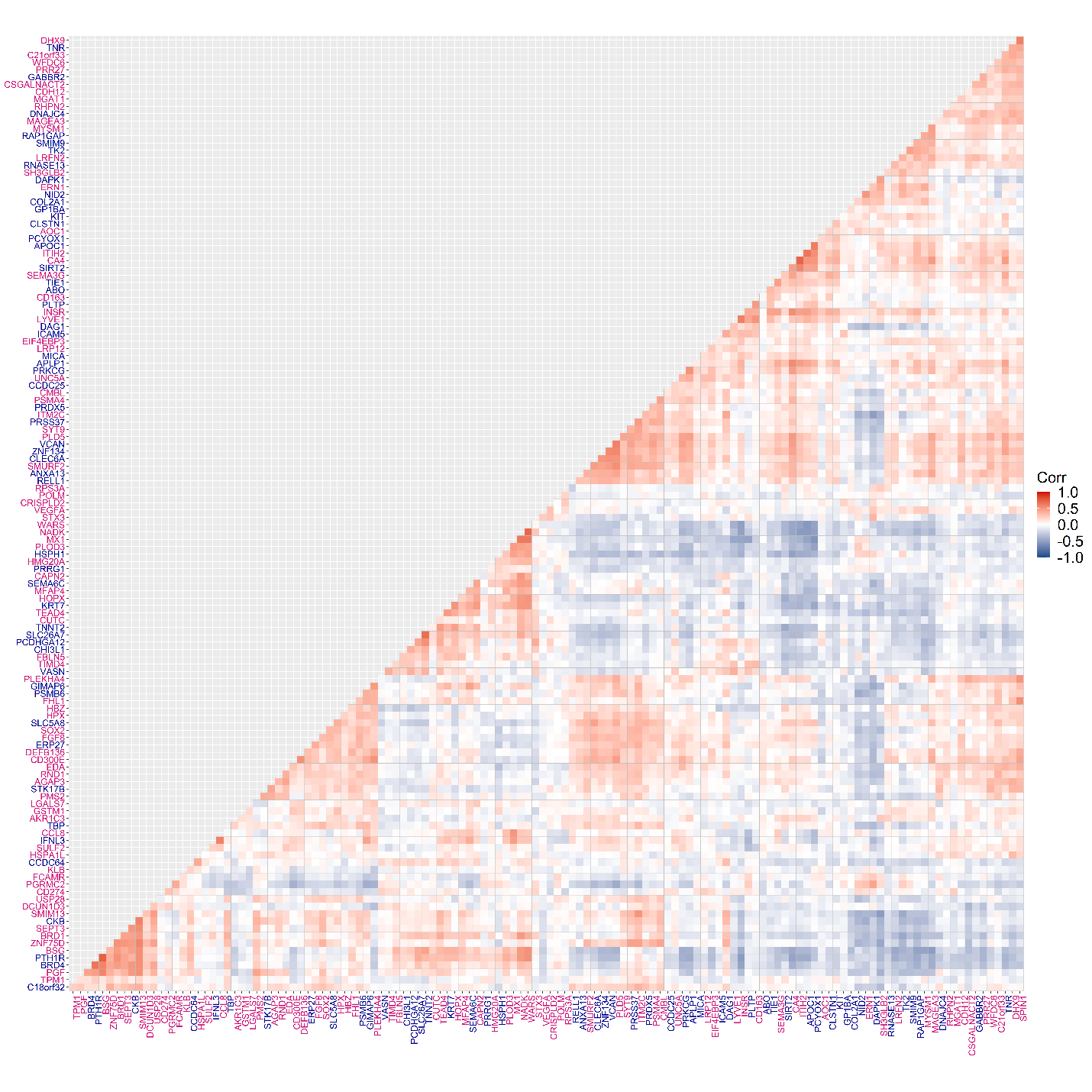

**Supplementary Figure 5 | Correlations of SOMAmer reagents selected by the protein model when predicting COVID-19 outcomes. a**, Spearman’s rank correlations between the 92 proteins within the protein model trained to predict the severe COVID-19 outcome. Plot shows a 92 x 92 heatmap containing 8464 total correlations with only the bottom portion of the heatmap being shown. Proteins were hierarchically clustered with 8372 correlations (98.9%) having Spearman's absolute ρ < 0.8. **b**, Spearman’s rank correlations between the 67 proteins within the protein model trained to predict the critical COVID-19 outcome. Plot shows a 67 x 67 heatmap containing 4489 total correlations with only the bottom portion of the heatmap being shown. Proteins were hierarchically clustered with 4422 correlations (98.5%) having Spearman's absolute ρ < 0.8.

**c**, Spearman’s rank correlations between the nonoverlapping proteins predicting severe (dark blue) and critical (pink) COVID-19 outcomes. Plot shows a 131 x 131 heatmap containing 17161 total correlations with only the bottom portion of the heatmap being shown. Proteins were hierarchically clustered with 17030 (99.2%) having Spearman’s absolute ρ < 0.8.
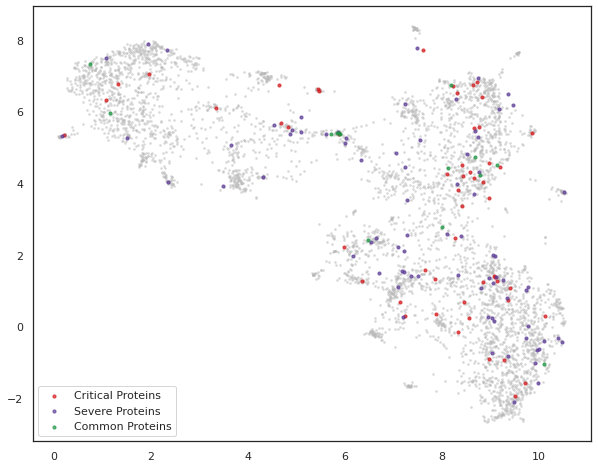

**Supplementary Figure 6 | Unsupervised clustering by uniform manifold approximation and projection (UMAP) on 4984 SOMAmer reagents in the BQC19 cohort.** Nonlinear dimensionality reduction of measured levels of 4984 SOMAmer reagents from the BQC19 cohort projected into a 2-dimensional space. Proteins selected by L1 regularized logistic regression for predicting critical COVID-19 using the protein model are shown in red and proteins selected for predicting severe COVID-19 are shown in purple. The 14 common proteins that were selected in protein models trained on both severe and critical COVID-19 are shown in green.
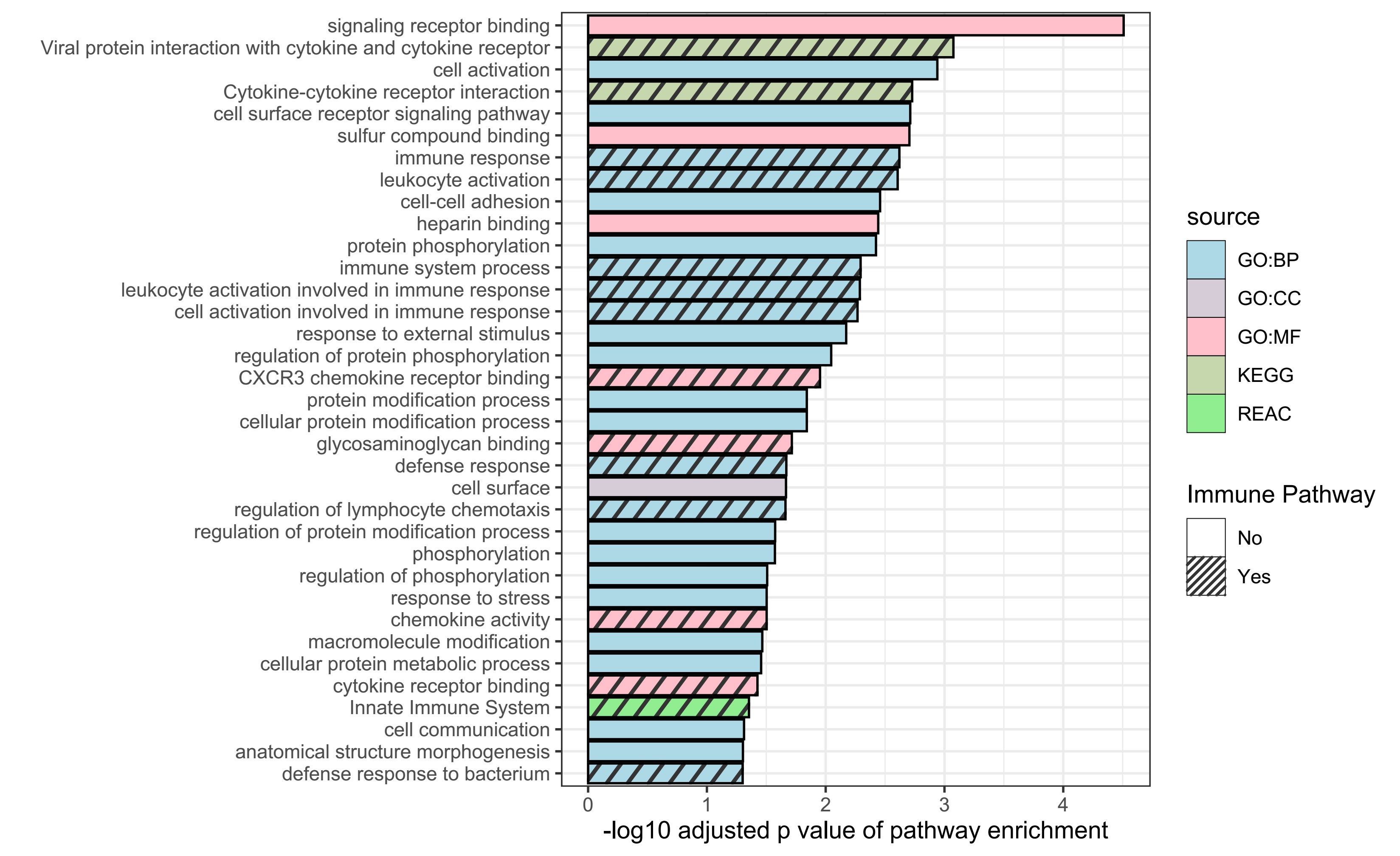

**Supplementary Figure 7 | Pathway analysis on common proteins best predicting severe and critical COVID-19 outcomes.** 32 proteins were common between proteins selected by LASSO that are predictive of critical COVID-19 plus their highly correlated proteins (N=96, Spearman's absolute ρ > 0.75) and proteins selected by LASSO that are predictive of severe COVID-19 plus their highly correlated proteins (N=171, Spearman's absolute ρ > 0.75).

Pathway enrichment was predicted by g:Profiler. Pathways with g:SCS adjusted P < 0.05 are listed. GO_BP: Gene Ontology biological process; GO_CC: Gene Ontology cellular component; GO_MF: Gene Ontology molecular function; KEGG: KEGG: Kyoto Encyclopedia of Genes and Genomes; REAC: Reactome Pathway Database. Diagonally patterned bars represent immune response pathways.

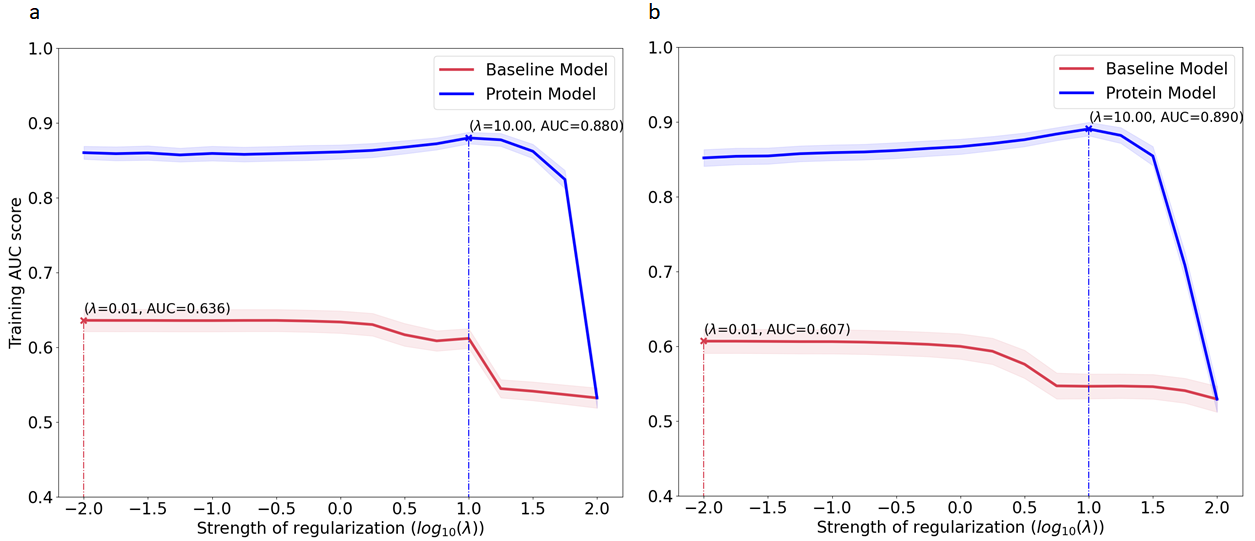
 c	d
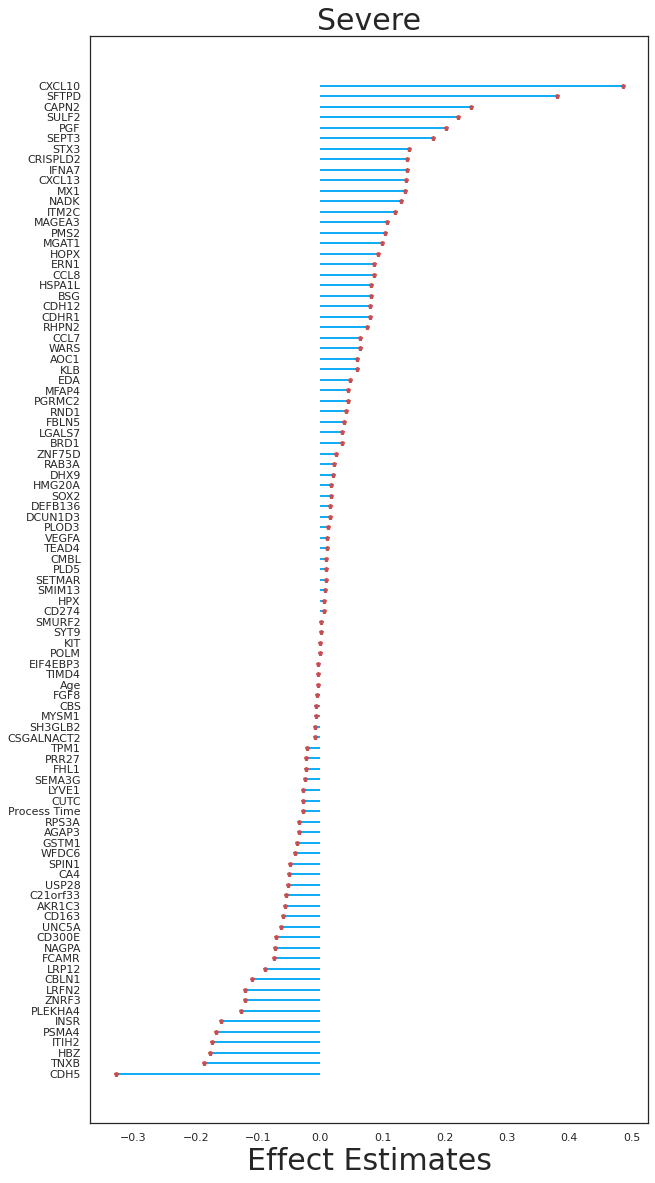

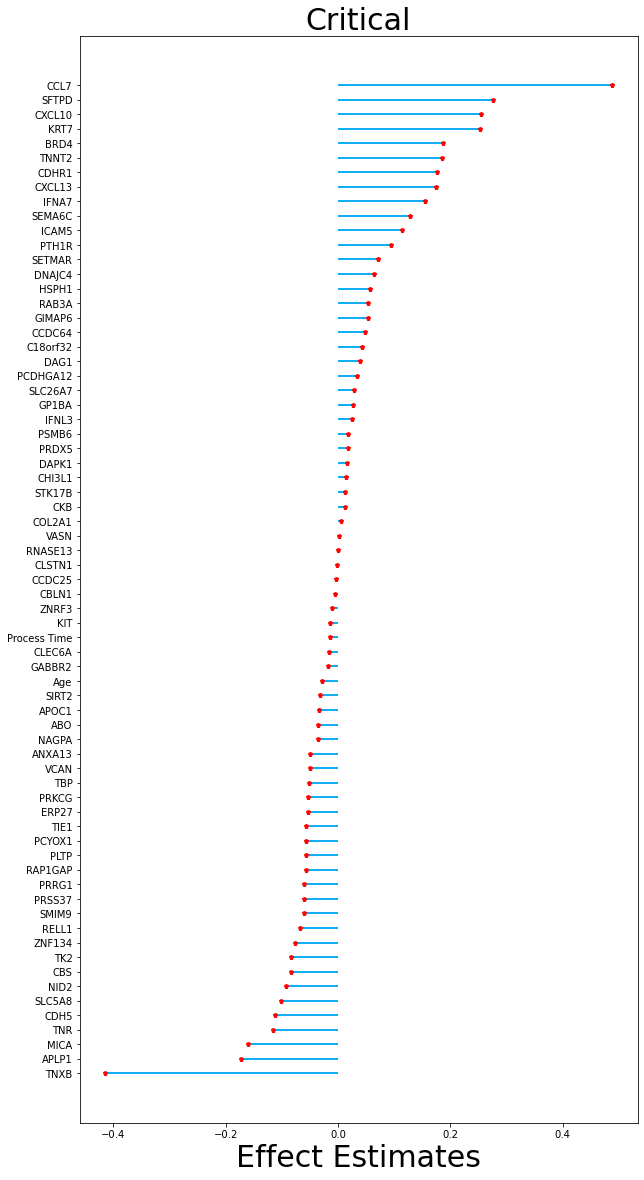

**Supplementary Figure 8 | Main Analysis plus 6 clinical risk factors.**

We trained a baseline model (red) with covariates age, sex, sample processing time, hospital site, and 6 clinical features as well as a protein model (blue) containing all baseline model covariates along with an additional 4984 SOMAmer reagents. The sample size used in this analysis was the entire BQC19 cohort comprising 417 patients.

Train AUC score as a function of regularization strength during L1 regularized logistic regression when training to predict **a,** severe COVID-19 and **b,** critical COVID-19. The baseline model was fitted with covariates age, sex, sample processing time, hospital site, and 6 clinical features (diabetes, chronic obstructive pulmonary disease, chronic kidney disease, congestive heart failure, hypertension, liver disease). The protein model was fitted with all variables in the baseline model along with 4984 SOMAmer reagents. Each data point represents an AUC score averaged over 50 validation folds. Shaded areas represent 95% confidence intervals above and below the mean AUC. **c**, Absolute values of the 95 nonzero coefficients of the final trained L1 regularized logistic regression protein model for predicting severe COVID-19. The original data contained 4984 proteins and 4 variables age, sex, sample processing time, hospital site, smoking status along with 6 clinical features. A total of 93 proteins remained within the model along with age and sample processing time. The model was trained on the entire training set using lambda = 10.0 (log_10_ lambda = 1.0) which was the best lambda value found from the hyperparameter search. **d**, Absolute values of the 69 nonzero coefficients of the final trained L1 regularized logistic regression protein model for predicting critical COVID-19. The original data contained 4984 proteins and 4 variables age, sex, sample processing time, hospital site, along with 6 clinical features. A total of 67 proteins remained within the model along with age and sample processing time. The model was trained on the entire training set using lambda = 10.00 (log_10_ lambda = 1.0) which was the best lambda value found from the hyperparameter search.

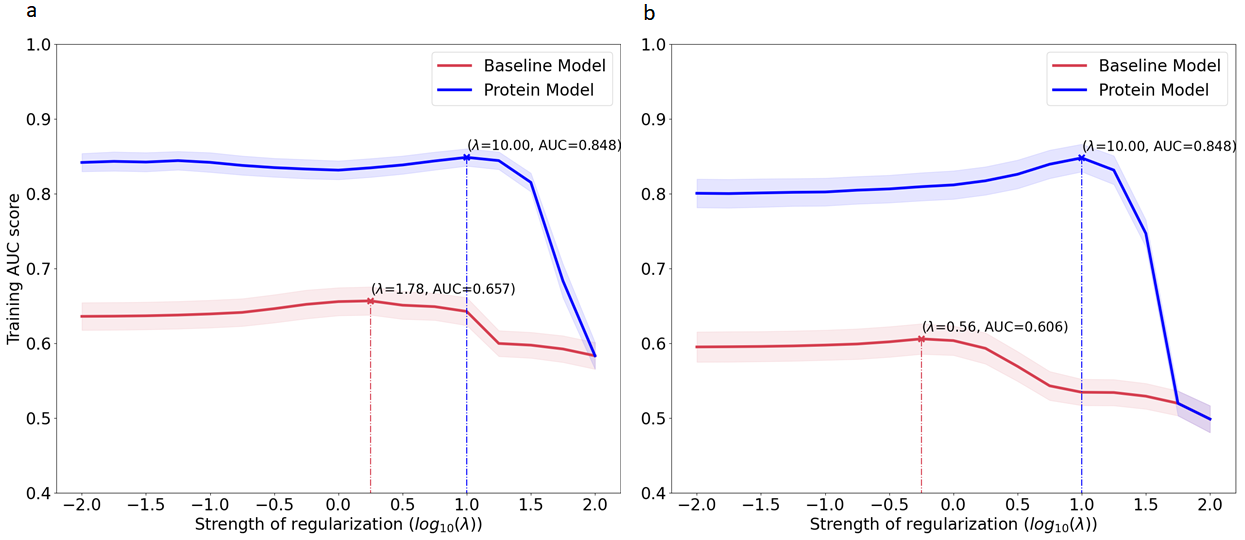
c	d
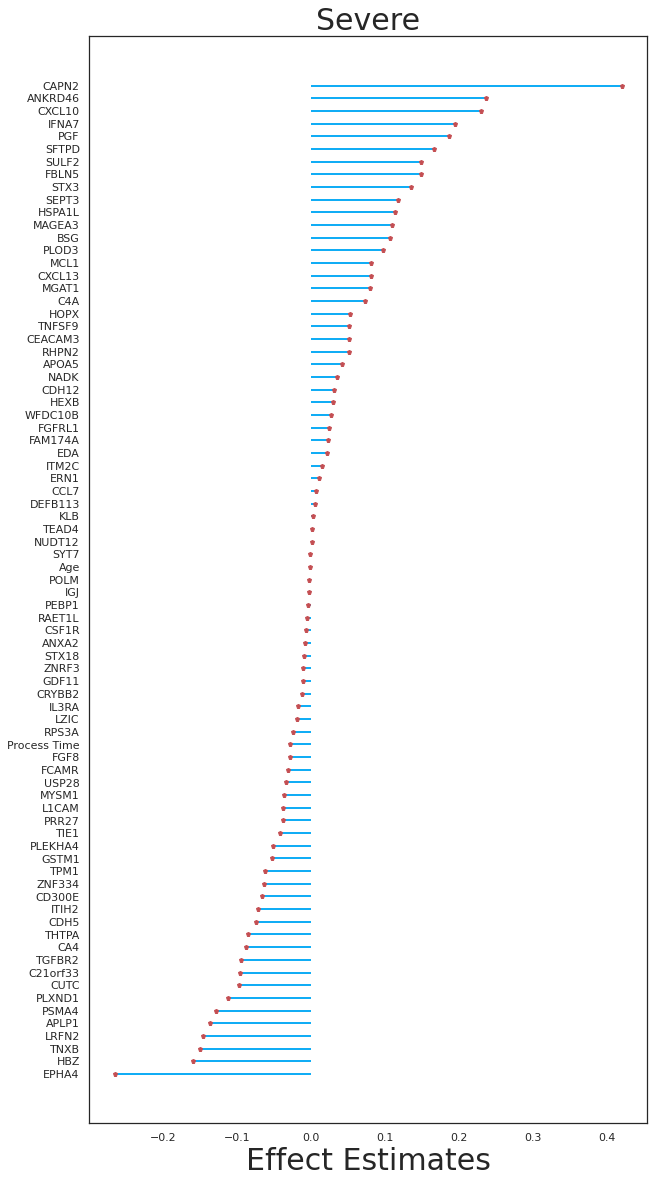

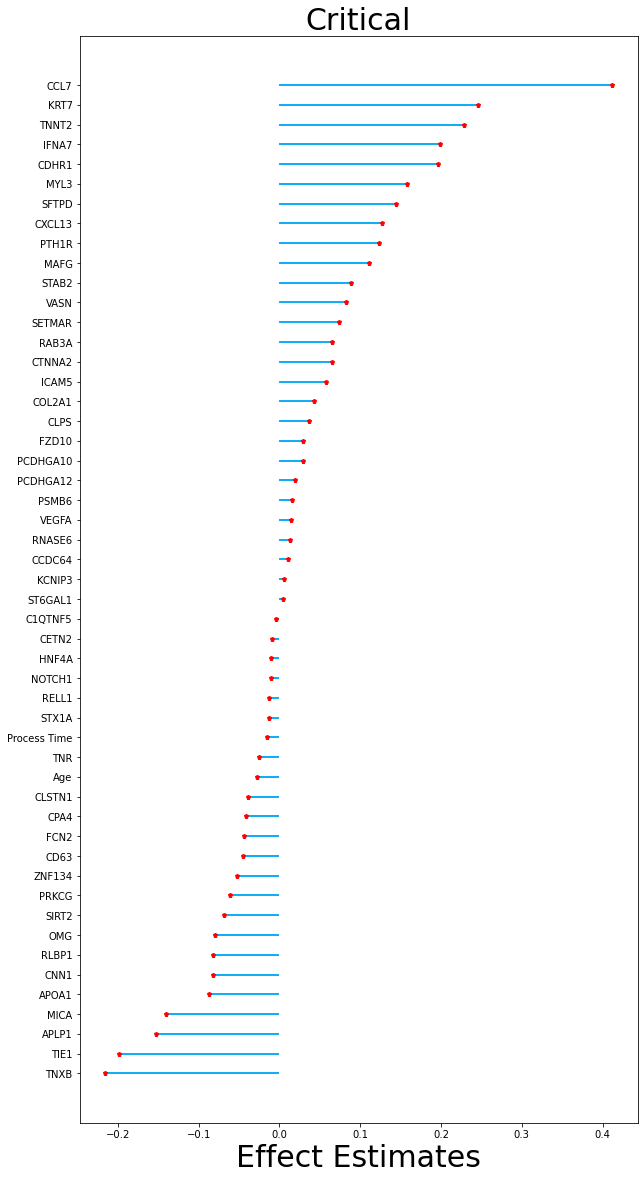

**Supplementary Figure 9 | Main Analysis plus 7 clinical risk factors.**

We trained a baseline model (red) with covariates age, sex, sample processing time, hospital site (JGH only), 6 clinical features, and smoking status as well as a protein model (blue) containing all baseline model covariates along with an additional 4984 SOMAmer reagents. Due to the smoking status variable missing from the CHUM data, the sample size was reduced from 417 samples down to 312 samples for the analysis.

Training AUC score as a function of regularization strength during L1 regularized logistic regression when trained to predict **a**, severe COVID-19. **b,** critical COVID-19. The baseline model was fitted with covariates age, sex, sample processing time, hospital site, smoking, and 6 clinical features (diabetes, chronic obstructive pulmonary disease, chronic kidney disease, congestive heart failure, hypertension, liver disease). The protein model was fitted with all variables in the baseline model along with 4984 SOMAmer reagents. Each data point represents an AUC score averaged over 50 validation folds. Shaded areas represent 95% confidence intervals above and below the mean AUC. **c**, Absolute values of the 79 nonzero coefficients of the final trained L1 regularized logistic regression protein model for predicting severe COVID-19. The original data contained 4984 proteins and 4 variables age, sex, sample processing time, hospital site, smoking status along with 6 clinical features. A total of 77 proteins remained within the model along with age and sample processing time. The model was trained on the entire training set using lambda = 10.00 (log_10_ lambda = 1.0) which was the best lambda value found from the hyperparameter search. **d**, Absolute values of the 51 nonzero coefficients of the trained L1 regularized logistic regression protein model for predicting critical COVID-19. The original data contained 4984 proteins and 4 variables age, sex, sample processing time, hospital site, smoking status along with 6 clinical features. A total of 49 proteins remained within the model along with age and sample processing time. The model was trained on the entire training set using lambda = 10.00 (log_10_ lambda = 1.0) which was the best lambda value found from the hyperparameter search.
Supplementary Notes

*Using elastic net to select uncorrelated proteins*

For elastic net, we used elastic net regularized logistic regression (linear combination of L1 and L2 regularization). To implement this elastic net, we used the “LogisticRegression” module from sci-kit learn with penalty set to “elasticnet”. The elastic net penalty combines both the L1 norm and the L2 norm penalties. The L1 regularization term forces coefficients to be zero and in effect performs feature selection. The L2 regularization term does not force coefficients to have a null effect but instead reduces their magnitude, resulting in a model with a greater number of variables but many with small effect estimates. Therefore, the combination of both of these regularization terms gives rise to a more generalized form of the L1 regularized logistic regression model used in the main analysis. For model training, the first step requires tuning two hyperparameters. In addition to the hyperparameter lambda which controls the amount of L1 regularization as previously described for LASSO logistic regression, the hyperparameter “alpha” must also be tuned. In Sci-kit learn, this parameter is termed the “L1 ratio” and ranges between 0 and 1 while controlling the amount of L1 to L2 regularization. For example, if the L1 ratio is set to 1, this is in essence the L1 regularized logistic regression model. Setting the L1 ratio to the other extreme, 0, results in an L2 regularized logistic regression model which is termed Ridge Logistic Regression. The tuning of these two hyperparameters was also performed through cross-validation. For the lambda hyperparameter, we searched over the same range of values as that used in L1 regularized logistic regression in the main analysis. For the L1 ratio, we searched over 11 values: [0, 0.1, 0.2, 0.3, 0.4, 0.5, 0.6, 0.7, 0.8, 0.9, 1.0].

For the elastic net regularized logistic regression model, we used a tolerance parameter of 0.01 during training instead of 0.0001 used for L1 regularized logistic regression to speed up tuning. For reproducibility, the random seed was set to 0 for cross validation splits as well as weight initialization during model training. The training performance between L1 and elastic net logistic regression was generally similar, so we decided to use the L1 regularized logistic regression model to test on the external cohort due to it being faster to train and less complex than elastic net. We used the best hyperparameter lambda, selected from cross validation, to train the baseline and protein model using the entire BQC19 cohort. We pre-processed the training dataset by natural log transforming the protein levels due to protein levels varying considerably. The binary predictor variables sex and sample processing time were dummy encoded while the continuous variables age and sample processing time were left as is. All protein variables were standardized to a mean of zero and unit variance.

*Training and performance of models using elastic net*

We also performed elastic net regularized logistic regression which is a more generalized form of L1 regularized logistic regression as it combines both L1 and L2 penalty terms. Instead of directly setting protein coefficients to zero such as in LASSO, the elastic net contains an additional L2 penalty which allows some protein coefficients to remain nonzero but reduces their effect estimates (i.e., it reduces the size of the coefficients). Cross-validation was performed similarly to L1 regularized logistic regression but for elastic net, in addition to the lambda hyperparameter, we tuned an extra parameter termed the L1 ratio which determines the amount of L1 to L2 regularization to use. The cross-validation results are shown in (**Supplementary Figure 4**). The best hyperparameters produced training AUCs of 87.3% and 88.5% for the protein model when predicting severe COVID-19 and critical COVID-19, respectively (data not shown). For the baseline model, training AUC results for predicting severe COVID-19 and critical COVID-19 were 54.6% and 54.9%, respectively (data not shown). The training performance results for elastic net models were similar to LASSO models with the only improvement demonstrated by the protein model when predicting severe COVID-19 (LASSO training AUC 88.0% vs. elastic net training AUC 87.3%). Due to small differences between LASSO and elastic net performance, we did not pursue testing of the elastic net model in the external Mount Sinai cohort since it is more complex and requires more computational time to train.
